# Conformal Prediction and Venn-ABERS Calibration for Reliable Machine Learning-Based Prediction of Bacterial Infection Focus

**DOI:** 10.1101/2025.01.21.25320878

**Authors:** Jacob Bahnsen Schmidt, Karen Leth Nielsen, Dmytro Strunin, Nikolai Søren Kirkby, Jesper Qvist Thomassen, Steen Christian Rasmussen, Ruth Frikke-Schmidt, Frederik Boëtius Hertz, Allan Peter Engsig-Karup

## Abstract

Finding the focus of bacterial infections can be challenging, especially for hospitalised patients. Conventional microbiological diagnostic methods are either time consuming, expensive, or difficult to interpret due to contamination. The aim of this study was to apply machine learning (ML) to reliably predict the focus of bacterial infections. This study utilised a dataset including samples from 10,153 patients, collected from November 1, 2019, to June 3, 2023, at Rigshospitalet, Denmark. The dataset contains microbiological findings, biochemical data, and vital parameters. The dataset was analysed using ML. The ML-outputs were calibrated using Venn-ABERS calibration and model uncertainty was addressed using conformal risk control. The best performing model was the XGBoost model achieving a Log loss of 0.219 ± 0.050 (mean ± SD.) and an AUC of 0.93 ± 0.051. Combining the model with methods from the conformal prediction framework achieves predictive capabilities that surpasses similar studies, while also accounting for model uncertainty by providing statistically robust uncertainty estimates via calibrated probabilistic predictions.

## 1. Introduction

The global mortality rate from bacterial infections remains alarmingly high. A study estimated that 7.7 million deaths worldwide in 2019 were caused by 33 different genera and species of bacteria^1^. The mortality rate associated with these infections was 52.2 deaths per 100,000 population in high-income countries. Many of these deaths are due to delayed antibiotic therapy or inadequate treatment of infections, in many cases due to resistance of the causative bacteria^2^.

Clinical microbiological diagnostics provide clinicians with important information, such as the identity of the pathogen and its antimicrobial resistance profile for the treatment of infectious diseases. Today’s microbiological diagnostics primarily rely on phenotypic testing, which can be slow and time-consuming due to the need for bacterial growth^3,4^. This delay in diagnosis often leads to the use of broad-spectrum antibiotics. However, narrow-spectrum antibiotics are usually more effective against specific types of bacteria and have fewer side effects compared to broad-spectrum antibiotics^3^. Ensuring that the most effective antibiotics are administered as early as possible, which in turn improves patient outcomes and ultimately lowers the global mortality rate from bacterial infections.

Furthermore, reducing the unnecessary use of antibiotics and allowing for a faster transition from broad-spectrum antibiotics to narrow-spectrum antibiotics helps mitigate the emergence of multi-drug resistance and preserve the effectiveness of existing antibiotics. In addition to the early and accurate identification of the bacterial pathogen and its antimicrobial resistance profile, it is crucial to determine whether the isolated pathogen is truly causing the disease or is merely a contaminant from the clinical specimen. Clinicians must assess patient history, vital parameters, and laboratory test results to decide if the patient has an infection or is at risk for developing severe infections before administering antibiotics. If treatment is necessary, they must decide which antibiotic to use, whether it should be given orally or intravenously, and the appropriate dosage. Not only the identity of the pathogen and its resistance profile, but also the severity of the disease and the site of infection significantly influences the selection, effectiveness, and management of antibiotic therapy. Consequently, making treatment decisions and choosing the appropriate antibiotic can be complex, requiring clinical expertise and additional laboratory or imaging information. Recent antibiotic stewardship initiatives, such as the “Start smart then focus toolkit” from (www.gov.uk) attempt to guide clinicians in rational antibiotic use. Machine learning (ML) could be a valuable predictive tool, providing diagnosticians with decision support in the early stages of diagnosing bacterial infections.

Previous studies have shown promising results in predicting bacterial infections using data-driven ML approaches. A study by Marandi et al.^5^ focused on predicting bloodstream infection (BSI) primarily based on biochemical data. Their best performing model, LightGBM, achieved an AUC of 0.69, with a sensitivity of 0.57 and specificity of 0.70. Rawson et al.^6^ used support vector machines (SVM) to predict the onset of community-acquired bacterial infections within the first 72 hours of hospital admission. Using six commonly available blood parameters, they achieved an AUC of 0.84, with a sensitivity of 0.89 and specificity of 0.63 at a likelihood cut-off of 0.81. Slightly adjusting the cut-off to 0.82 resulted in a sensitivity of 0.44 and specificity of 0.93, indicating potential model uncertainty and calibration issues.

In this study, we aimed to develop a data-driven approach using classical ML-models combined with the conformal prediction framework^7^, to create a reliable, fast and cost-effective datadriven ML-model for supporting clinicians and diagnosticians in diagnosing bacterial infections. Classical ML-models such as XGBoost are algorithms that learn patterns in data to make predictions, similar to how a clinician might consider multiple risk factors when assessing a patient. The conformal prediction framework allows such models to provide a measure of confidence or uncertainty with each prediction, helping to indicate how reliable the prediction is in a given case. This method addresses known issues related to model uncertainty in the field of ML, providing more robust and interpretable predictions. By leveraging electronic health records and biochemical data, our goal was to create a foundation for a future datadriven ML model that, by utilizing the conformal prediction framework, can offer accurate predictions with well-calibrated confidence levels, ultimately enhancing decision-making in clinical settings and improving patient outcomes.

## 2. Methods

### 2.1. Data Collection

This study utilised a retrospective cohort design, including patients of all age groups who had positive bacterial cultures sampled at Rigshospitalet (Copenhagen, Denmark) between November 1, 2019 and June 3, 2023.

Rigshospitalet serves as a tertiary hospital primarily for referral and highly specialised treatment. The dataset was produced using Danish electronic health records, i.e., patient journals, by combining data from the electronic patient journals (Sundhedsplatformen, Epic Systems Corporation) and the MADS database (Department of Microbiology Data Systems, in Danish Mikrobiologisk Afdelings Data System). Doublets, defined as subsequent isolates stemming from the same patient with the same outcome within 30 days, were removed from the dataset.

The dataset was compiled as a cross-sectional dataset to simplify the analysis while ensuring that only the most recent observations were used, allowing the model to be applicable to new cases without requiring access prior recorded data.Biochemical test results from blood samples and vital parameters were gathered up to seven days prior to the positive conventional cultures, to ensure that the model is trained on data relevant to the bacterial infection in question. The biochemical test results and vital parameters were aggregated using the mean on each day per patient. This resulted in a dataset consisting of 73 features (Table 1). 58 describing the patient’s biochemical test results, ten describing the patient’s vital parameters, two describing the patient’s age and sex, and three features gained from feature engineering. Furthermore, data transformations were applied to each feature to obtain Gaussian distributed features, as described in S6. For the purpose of supervised machine learning (ML), the dataset was labelled using a table describing the findings related to the bacterial isolates. Specifically, ML labels were generated using a combination of the sample origin and type of bacteria found in the sample (S15 shows the ML-label assigned to each combination of bacteria and origin).

**Table 1.** Table of summary statistics (Median, [IQR] and number of observations) grouped by the infection type (For binary variables only the number of observations is given). Variable describes the name of the feature in the dataset and description is a short description of the feature. Airway is airway infections; BSI is blood stream infections; and UTI is urinary tract infections. A full table describing all the variables in all infection focuses is available in S16.

| Variable | Description | Unit | Airways | BSI | UTI |
| --- | --- | --- | --- | --- | --- |
| Sex | Patient sex | Binary | Male 779<br>Female 449 | Male 1774<br>Female 1194 | Male 4069<br>Female 4069 |
| Age | Patient age | Years | 59<br>[44.00, 69.00]<br>1228 | 62<br>[42.00, 71.00]<br>2968 | 67<br>[53.00, 76.00]<br>11430 |
| ALB | Albumin | g/L | 26<br>[21.00, 32.00]<br>700 | 26.5<br>[22.00, 31.00]<br>1405 | 31<br>[26.00, 36.00]<br>6517 |
| BASO | Basophilocyte count | 10<sup>9</sup>/L | 0.03<br>[0.01, 0.06]<br>510 | 0.03<br>[0.01, 0.05]<br>911 | 0.03<br>[0.01, 0.06]<br>5711 |
| BILI | Bilirubin | μmol/L | 10<br>[6.00, 20.00]<br>563 | 12<br>[7.00, 23.61]<br>1499 | 8<br>[5.00, 14.00]<br>3915 |
| BILIKON | Conjugated bilirubin | μmol/L | 101<br>[8.10, 194.75]<br>4 | 13.5<br>[9.00, 18.75]<br>30 | 2<br>[2.00, 5.25]<br>104 |
| CARB | Carbamide | mmol/L | 8.5<br>[5.50, 14.00]<br>947 | 8.95<br>[5.70, 15.30]<br>1499 | 6.9<br>[4.50, 11.10]<br>5925 |
| CREA | Creatinine | μmol/L | 81<br>[55.50, 128.50]<br>1157 | 82<br>[58.00, 136.00]<br>2689 | 72<br>[54.00, 107.00]<br>10415 |
| CRP | C-reactive protein | mg/L | 57<br>[17.00, 139.00]<br>1165 | 45<br>[11.00, 116.88]<br>2602 | 21<br>[6.00, 58.00]<br>9505 |
| DiastolicBP | Diastolic blood pressure | mmHg | 60.41<br>[54.57, 68.82]<br>1149 | 66.5<br>[57.39, 75.00]<br>2698 | 68.83<br>[60.58, 77.75]<br>9999 |
| EWS | Early warning score | Integer | 2.67<br>[1.00, 4.36]<br>342 | 1.33<br>[0.50, 3.00]<br>1789 | 1<br>[0.00, 2.00]<br>7231 |
| HB | Hemoglobin | mmol/L | 6.2<br>[5.40, 7.20]<br>1201 | 6.1<br>[5.40, 7.05]<br>2833 | 6.9<br>[5.90, 7.90]<br>10939 |
| HDL | High density lipoprotein | mmol/L | 1.02<br>[0.85, 1.40]<br>28 | 1.01<br>[0.67, 1.35]<br>70 | 1.31<br>[1.04, 1.67]<br>972 |
| HR | Heart rate | min<sup>-1</sup> | 88<br>[77.01, 100.54]<br>1178 | 89.5<br>[78.50, 101.87]<br>2741 | 81.73<br>[71.50, 93.50]<br>10145 |
| LDH | Lactate dehydrogenase | U/L | 295<br>[205.00, 452.00]<br>625 | 240<br>[182.00, 359.00]<br>1233 | 221<br>[182.00, 286.00]<br>4010 |
| LEU | Leukocyte count | 10<sup>9</sup>/L | 10.9<br>[7.60, 14.83]<br>1176 | 8.5<br>[4.10, 13.00]<br>2690 | 9.1<br>[6.70, 12.30]<br>9946 |
| LYMFO | Lymphocyte count | 10<sup>9</sup>/L | 1.23<br>[0.71, 1.81]<br>509 | 0.95<br>[0.51, 1.50]<br>910 | 1.37<br>[0.93, 1.91]<br>5712 |
| NEUTRO | Neutrophil count | 10<sup>9</sup>/L | 7.9<br>[5.47, 11.00]<br>495 | 7.13<br>[4.31, 11.00]<br>889 | 6.42<br>[4.48, 9.14]<br>5573 |
| O2Therapy | Oxygen therapy | L/min | 35.15<br>[3.50, 50.87]<br>1113 | 0<br>[0.00, 25.86]<br>2104 | 0<br>[0.00, 2.67]<br>7918 |
| PROCAL | Procalcitonin | µg/L | 0.66<br>[0.22, 2.59]<br>247 | 0.81<br>[0.27, 2.94]<br>243 | 0.29<br>[0.10, 0.69]<br>344 |
| Saturation | Oxygen saturation | % | 96.37<br>[94.72, 97.80]<br>1171 | 97.5<br>[96.17, 99.00]<br>2631 | 97.5<br>[96.00, 98.67]<br>9745 |
| SystolicBP | Systolic blood pressure | mmHg | 119.32<br>[106.00, 132.82]<br>1158 | 122<br>[107.33, 136.50]<br>2717 | 128.87<br>[115.25, 143.92]<br>10028 |
| Temperature | Body temperature | °C | 37.15<br>[36.66, 37.64]<br>1145 | 37.1<br>[36.60, 37.75]<br>2650 | 36.95<br>[36.55, 37.41]<br>9624 |
| eGFR | Estimated Glomerular Filtration Rate | mL/min | 77<br>[45.00, 90.00]<br>1061 | 71<br>[38.00, 90.00]<br>2452 | 80<br>[50.00, 90.00]<br>9870 |
| Weight | Weight | kg | 80<br>[63.88, 95.00]<br>599 | 72.8<br>[57.50, 86.60]<br>1329 | 71.33<br>[58.57, 84.25]<br>4555 |

The labels in this study included three foci of infection: airway infections (airways), blood stream infections (BSI), urine tract infections (UTI). For each focus of infection, we categorized bacteria into two groups: (1) typical pathogens (hereafter referred to as “pathogens”) and (2) organisms that are usually not pathogens or are common contaminants (hereafter referred to as “PS thus PSBSI, PSUTI, PSairways”). This classification was based on recent publications, adjusted to local guidelines, and considered the setting of a referral hospital. We used the following sources to inform our categorization^8–10^.

This approach allowed us to differentiate between organisms likely to cause true infections and those more likely to represent contamination or colonization in this specific context.

Due to the species included for sequencing, the class PSBSI was excluded from the analysis as this class only included 11 positive blood samples. Figure 1 provides an overview of the study design.

**Figure 1.**
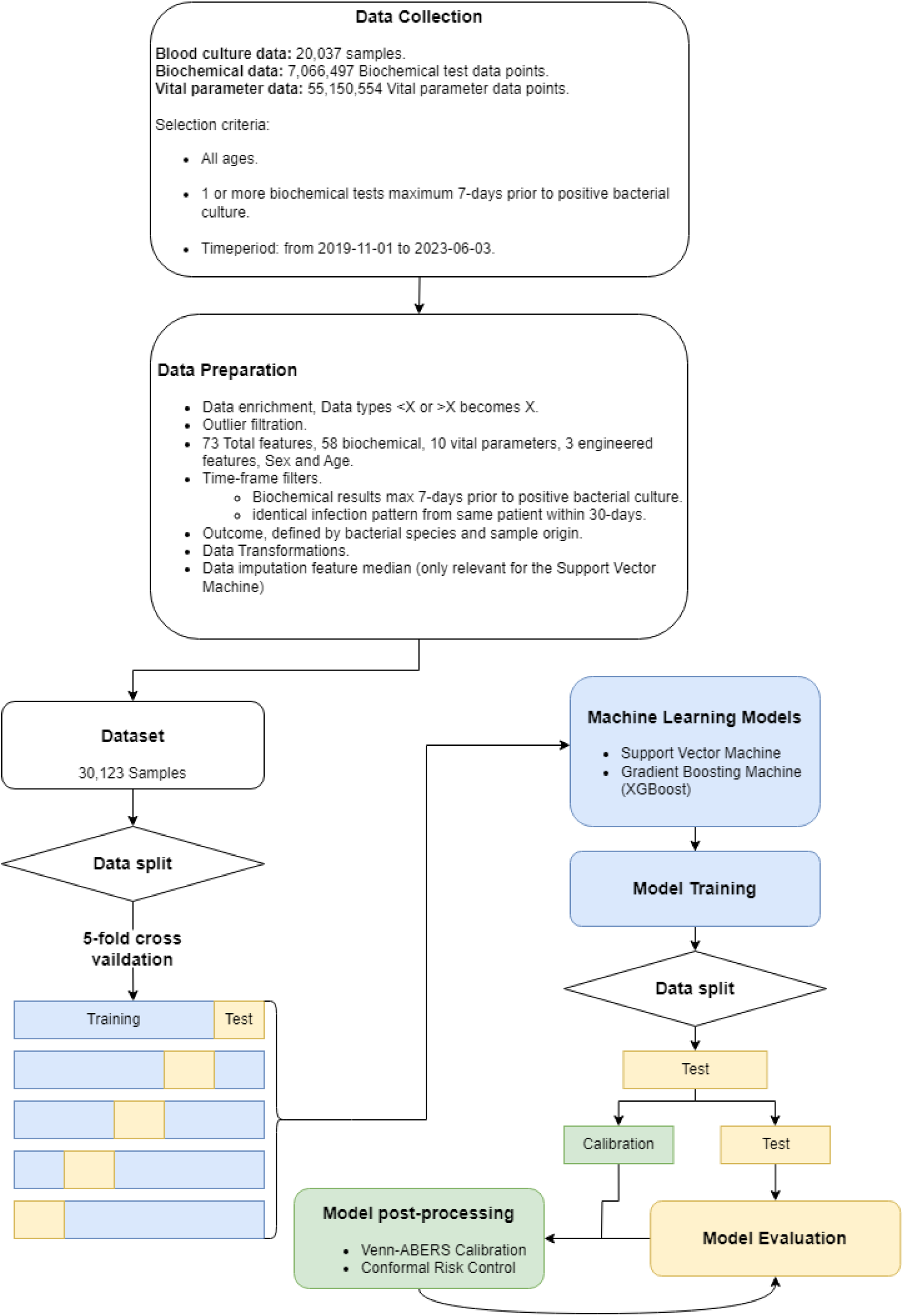
Overview of the Machine Learning pipeline used in this study from data collection to model evaluation.

### 2.2. Machine Learning

The dataset includes patients with a positive bacterial culture isolated from a wide range of body sites. The ML problem was framed as a seven dimensional one-vs-rest binary classification task. The goal was to create a model that could distinguish between seven different classes (different foci of infections and colonizations), each of them treated separately in a one-vs-rest approach, where the model learns to identify a particular focus of infection/colonization (the “one”) and distinguish it from the combined group of the remaining foci of infection/colonization (the “rest”). The ML-models were evaluated using 5-fold cross-validation, with an 80-20% split between training and test sets.

This study tested the use of a Support Vector Machine^11^ (SVM) and a Gradient Boosting Machine (GBM). These were implemented using Scikit-learn version 1.3.2 and python 3.8 - AzureML. Additionally, the XGBoost classifier^12^ version 1.3.3 was used, which is a scalable tree boosting algorithm building on the ideas of Friedman 2001^13^. XGBoost is an ensemble method that utilises a forest of simple classification trees. And can handle missing data, so no further preprocessing was necessary when training the XGBoost model. The model was optimised using a small grid search (sklearn’s GridSearchCV) and the optimal hyperparameters was found to be the default settings of the XGBoost model (for information on the SVM setup see S3).

Generally, ML-methods struggle with addressing model uncertainty. To tackle this issue, this study employed methods from the conformal prediction framework^7,14^, using a calibration dataset of 1000 observations. First the probabilistic model outputs were calibrated using the Venn-ABERS (VA) calibration technique^15^. VA calibration aims to adjust the probabilistic model outputs, to better match the ground truth.

In other words, if the model outputs a 42% probability of airway infection, this should accurately reflect the real-world likelihood. VA calibration uses a two-fold isotonic regression: one describes the probability of belonging to the class, and the other describes the probability of not belonging to the class. These regression models create a prediction interval (*p*0*, p*1) for an observation belonging to the positive class, with a guarantee that the true probability lies within this interval^16^. The adjusted probability outputs are calculated using the two isotonic regression models ^17^ and the formula in Eq. (1).

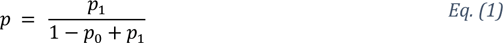

In practice, it is desirable to produce a model that can provide uncertainty guaranties. Therefore, this study employed the Conformal Risk Control (CRC) procedure from the conformal prediction framework^18^. CRC is a technique used to ensure reliable predictions that contain most of the true labels, while maintaining a user-specified error rate. In other words, CRC aims at creating prediction sets that are both accurate and meaningful.

The intuition behind CRC can be described in a handful of steps^14,18^. First the user chooses an acceptable error rate called *α*. This should be thought of as the maximum error rate that the user can tolerate to accept the ML prediction in a given application. Secondly, prediction sets for new observations are generated, ideally including the true labels of the observations. The third step is the balancing act, which ensures that the average error of the prediction sets, generated by a suitably chosen loss function is less than or equal to the chosen error rate. In the context of multi-label classification, as in this study, where each patient can have multiple focuses of infection, the goal is to produce a classifier that can assign accurate probabilities to each focus of infection and then choose thresholds, such that only predictions or focuses of infection with adequately high probability scores are included in the prediction set. This is an important part of the CRC procedure and essentially helps keeping the prediction sets small and meaningful, such that only the most likely outcomes are included in the prediction set.

Intuitively this also means that the size of the prediction sets wary with the difficulty of the new observation. If a new observation is diagnostically easy, then the resulting prediction sets will be small. However, if a new observation is difficult, then the prediction sets will be comparatively larger. Put differently, the model signals uncertainty in a human like fashion by generating prediction sets of different sizes based on how certain the model is.

The CRC framework requires a user chosen loss function. This study used the False negative rate (FNR) as the loss function, to control the sensitivity, setting the user specified error rate *α* = 0.1, which essentially ensures a sensitivity of at least 90% on average. For a more thorough and technical description of the mathematical implementation of CRC see S4.

This study has also assumed a random pool based active learning^19^ (AL) strategy for the sampling in order to estimate how much data is required to train similar models in the future.

We also performed an ablation study as part of a sensitivity analysis, removing all hematopoietic patients and patients with (*LEU* < 1 ⋅ 10^9^/*L*) from the dataset. A low leucocyte count may associate with increased risk of BSI, especially among patients with cancer or hematopoietic stem cell transplantation recipients and the presence of this population in our dataset may impact the model.

### 2.3. Model Evaluation

The models were evaluated using area under the receiver operating characteristics curve (AUC), as well as precision, sensitivity, specificity, F1-score, and multi class Brier score. Finally, this study examined the best performing model using Shapley additive explanations (SHAP). SHAP is a model agnostic tool that uses game theory to interpret the output of machine learning models^20^, that can be used with caution for interpretability^21^.

## 3. Results

This study included 10,153 hospitalized patients from both sexes and all age groups and from whom a bacterial isolate was recovered and processed at the Department of Clinical Microbiology (DCM) at Rigshospitalet between November 2019 and June 2023. From these 10,153 patients, 20,037 positive bacterial isolates of a diverse array of species (listed in S15) were processed at the DCM (on average 1.97 isolates per patient) and paired with 30,123 sets of biochemical test results. From these biochemical test results, 6,138 were related to isolates from the respiratory tract, 2,968 related to the blood stream, 13,052 related to the urinary tract and 9,922 related to other body sites.

The patients were categorized into groups based on the sample site of the clinical isolate. The first group comprised patients with a clinically apparent infection of the respiratory tract (1,228), the second with a blood stream infection (2,968) and the third with a urinary tract infection (11,430). There were 55 patients who were categorized as having a UTI as well as a respiratory tract infection, 364 patients were categorized with both a BSI and a UTI and 28 patients with simultaneous BSI and respiratory tract infection.

There were also patients from whom bacterial organisms that are usually not pathogens or are common contaminant were recovered from the respiratory tract (4,910), the urinary tract (1,622) or the blood stream (11). These patients were categorized into the three patient groups of airway, urinary tract and blood stream contamination / colonization. The group of BSI contaminations was however removed due to data sparsity. An overview of the patient group sizes is outlined in Figure 2, as an Euler diagram and a table, describing both the absolute number and overlaps in each of the 6 resulting infection classes.

**Figure 2.**
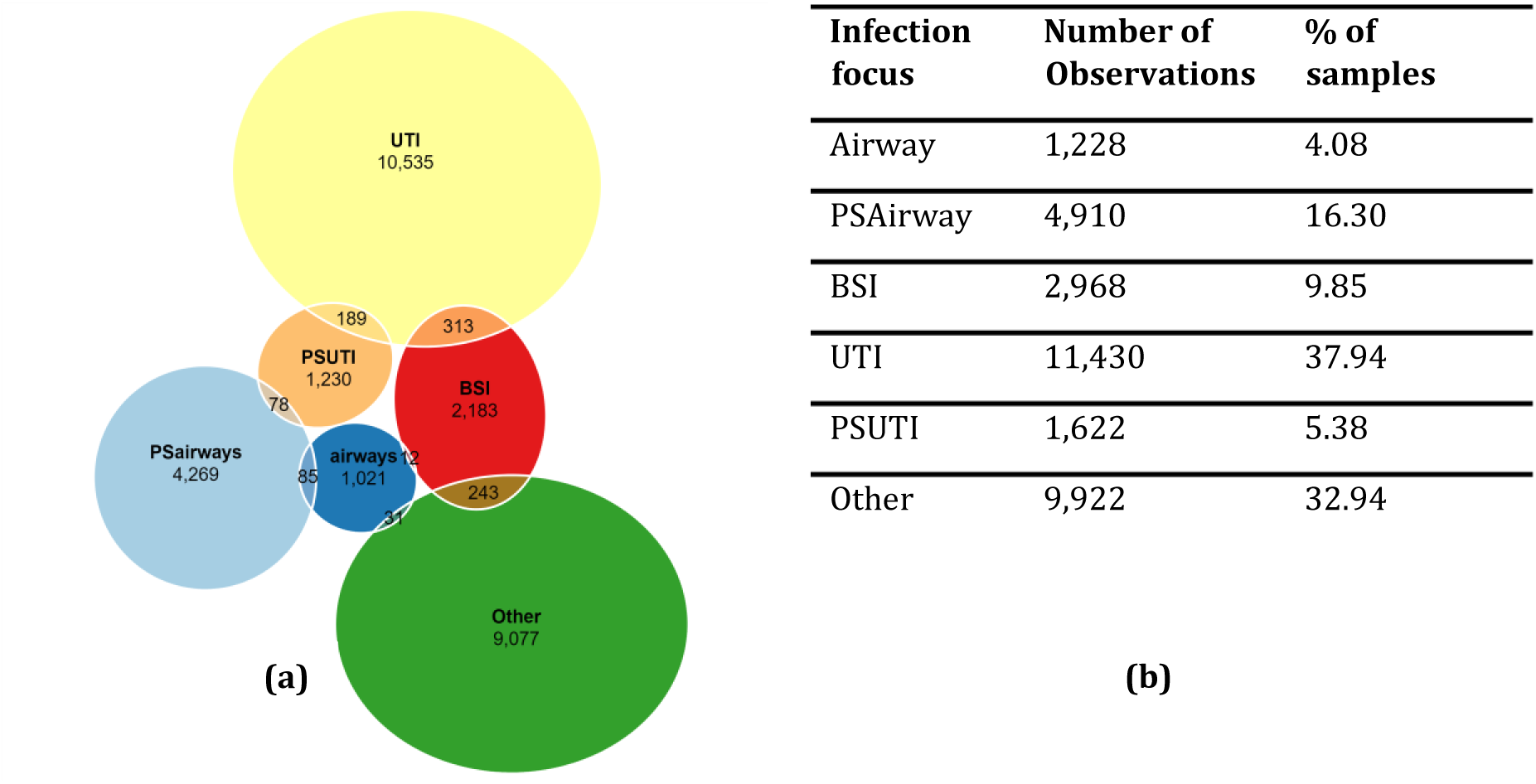
(a) shows an overview of the dataset, as an Euler diagram, where each ellipse represents an infection focus. The Euler diagram does not show all intersections between infection focuses, but rather the most important ones based on correlation between groups. The Euler diagram is fitted to the dataset using the eulerr R library. Figure 2(b) shows a summary of the dataset, showing the number of observations in each infection category. Airway includes airway infections, BSI is blood stream infections, UTI is urine tract infections, Other is an ensemble of other types of infection. PS denotes organisms that are usually not pathogens and common contaminants. This term is used to define observations that potentially could be contamination rather than infection. It should also be noted that the column % of samples sum to more than 100%, as patients can have multiple focuses of infection.

In the next step, 73 features were used as input for a model to predict the patient’s site of infection (respiratory tract, bloodstream, or urinary tract). These features included 58 biochemical test results, ten vital parameters, age, sex, and three features obtained through feature engineering (Table 1).

An overview of the model performance is presented in Table 2. The XGBoost model achieved the highest AUC and F1-score (mean ± std.) of 0.93 ± 0.051 and 0.71 ± 0.053 respectively. XGBoost outperforms the SVM on nearly all performance metrics. The sensitivity of the naïve XGBoost model was 0.63 ± 0.101 (mean ± std.), meaning it correctly predicted the label in 63% of the positive cases. The precision of the XGBoost model was 0.83 ± 0.094, indicating that if the model makes ten predictions, roughly eight will be correct. Additionally, the XGBoost model had a lower multi-class Brier score (mean ± std.) of 0.389 ± 0.103 and a Log loss of 0.219 ± 0.050, suggesting better calibration of probabilistic model outputs compared to the SVM. Applying the VA calibration substantially improved the F1-score for both models, indicating a better overall balance between precision and sensitivity. Using the CRC method achieved the desired sensitivity of at least 90%, but this increased sensitivity came with a trade-off in precision and specificity.

**Table 2.** Comparison table, showing the mean (std) micro averaged performance metrics across the 5-fold cross validation for each model, in both their naïve and VA calibrated states, as well as the model performance when the output have been optimized to satisfy the conformal risk control guarantee with an α = 0.1. PPV: Positive Predictive Value (Precision), SEN: Sensitivity, SPC: Specificity, F1: F1-score (measure of predictive performance – harmonic mean between PPV and SPC), MBS: Multi-class brier score (measure the accuracy of probabilistic predictions, lower score is better), AUC: area under the receiver operating characteristics curve, CPU TT: mean CPU time during training in minutes (std. in seconds), SVM: support vector machine, VA: Venn-ABERS calibration, CRC: conformal risk control, XGB: XGBoost, p-value: p-value for the test of difference in naïve models calculated using a permutation test with 1000 permutations.

| Metric \ Model | Naïve SVM | VA SVM | CRC-VA SVM | Naïve XGB | VA XGB | CRC-VA XGB | p-value (SVM vs. XGB) |
| --- | --- | --- | --- | --- | --- | --- | --- |
| **Micro averages** |  |  |  |  |  |  |  |
| PPV | **0.84**<br>(0.045) | 0.73<br>(0.045) | 0.44<br>(0.036) | 0.83<br>(0.094) | 0.79<br>(0.088) | 0.61<br>(0.100) | <0.001 |
| SEN | 0.37<br>(0.035) | 0.57<br>(0.092) | **0.90**<br>(0.015) | 0.63<br>(0.101) | 0.70<br>(0.127) | **0.90**<br>(0.013) | <0.001 |
| SPC | **0.94**<br>(0.008) | 0.90<br>(0.007) | 0.58<br>(0.074) | **0.94**<br>(0.022) | 0.92<br>(0.020) | 0.78<br>(0.121) | 0.973 |
| Log loss | 0.292<br>(0.024) | 0.271<br>(0.034) | .. | 0.219<br>(0.050) | **0.205**<br>(0.056) | .. | <0.001 |
| F1-score | 0.46<br>(0.044) | 0.63<br>(0.080) | 0.57<br>(0.034) | 0.67<br>(0.247) | **0.74**<br>(0.117) | 0.71<br>(0.087) | <0.001 |
| MBS | 0.523<br>(0.053) | 0.492<br>(0.068) | .. | 0.389<br>(0.103) | **0.366**<br>(0.114) | .. | <0.001 |
| AUC | 0.87<br>(0.045) | .. | .. | **0.93**<br>(0.051) | .. | .. | <0.001 |
| CPU TT | 9.92<br>(20.53) | .. | .. | **1.48**<br>(1.73) | .. | .. | .. |
| **Macro averages** |  |  |  |  |  |  |  |
| PPV | **0.89**<br>(0.079) | 0.73<br>(0.011) | 0.31<br>(0.183) | 0.84<br>(0.024) | 0.77<br>(0.040) | 0.52<br>(0.120) | <0.001 |
| SEN | 0.23<br>(0.204) | 0.51<br>(0.097) | **0.90**<br>(0.008) | 0.57<br>(0.105) | 0.67<br>(0.071) | **0.90**<br>(0.015) | <0.001 |
| SPC | **0.97**<br>(0.044) | 0.94<br>(0.060) | 0.59<br>(0.047) | **0.97**<br>(0.039) | 0.96<br>(0.044) | 0.80<br>(0.044) | 0.173 |
| F1-score | 0.31<br>(0.210) | 0.58<br>(0.066) | 0.42<br>(0.198) | 0.67<br>(0.069) | **0.71**<br>(0.053) | 0.63<br>(0.105) | <0.001 |
| **airways** |  |  |  |  |  |  |  |
| PPV | **0.88**<br>(0.171) | 0.71<br>(0.126) | 0.11<br>(0.030) | 0.83<br>(0.221) | 0.68<br>(0.345) | 0.43<br>(0.224) | <0.001 |
| SEN | 0.49<br>(0.224) | 0.49<br>(0.224) | **0.91**<br>(0.052) | 0.57<br>(0.228) | 0.63<br>(0.320) | 0.90<br>(0.023) | <0.001 |
| SPC | 0.99<br>(0.005) | 0.99<br>(0.005) | 0.66<br>(0.096) | **1.00**<br>(0.002) | **1.00**<br>(0.003) | 0.86<br>(0.204) | <0.001 |
| Log loss | 0.111<br>(0.020) | 0.093<br>(0.024) | .. | 0.069<br>(0.038) | **0.066**<br>(0.032) | .. | <0.001 |
| F1-score | 0.55<br>(0.214) | 0.55<br>(0.214) | 0.19<br>(0.048) | **0.67**<br>(0.247) | 0.65<br>(0.327) | 0.54<br>(0.235) | <0.001 |
| MBS | 0.026<br>(0.005) | .. | .. | **0.017**<br>**(0.010)** | .. | .. | <0.001 |
| AUC | 0.91<br>(0.053) | .. | .. | **0.95**<br>**(0.047)** | .. | .. | <0.001 |
| **PSairways** |  |  |  |  |  |  |  |
| PPV | **0.92**<br>**(0.046)** | 0.74<br>(0.074) | 0.26<br>(0.015) | 0.85<br>(0.120) | 0.78<br>(0.102) | 0.52<br>(0.126) | <0.001 |
| SEN | 0.14<br>(0.034) | 0.47<br>(0.114) | **0.90**<br>**(0.040)** | 0.51<br>(0.133) | 0.62<br>(0.169) | 0.89<br>(0.021) | <0.001 |
| SPC | **1.00**<br>**(0.001)** | 0.97<br>(0.007) | 0.52<br>(0.045) | 0.99<br>(0.008) | 0.97<br>(0.008) | 0.82<br>(0.115) | <0.001 |
| Log loss | 0.312<br>(0.028) | 0.286<br>(0.041) | .. | 0.238<br>(0.051) | **0.219**<br>**(0.064)** | .. | <0.001 |
| F1-score | 0.25<br>(0.055) | 0.57<br>(0.103) | 0.41<br>(0.019) | 0.63<br>(0.143) | **0.69**<br>**(0.149)** | 0.65<br>(0.114) | <0.001 |
| MBS | 0.091<br>(0.012) | .. | .. | **0.069**<br>**(0.019)** | .. | .. | <0.001 |
| AUC | 0.86<br>(0.044) | .. | .. | **0.92**<br>**(0.054)** | .. | .. | <0.001 |
| **BSI** |  |  |  |  |  |  |  |
| PPV | **0.94**<br>**(0.054)** | 0.73<br>(0.104) | 0.22<br>(0.111) | 0.84<br>(0.161) | 0.79<br>(0.137) | 0.42<br>(0.137) | <0.001 |
| SEN | 0.08<br>(0.028) | 0.42<br>(0.152) | 0.88<br>(0.068) | 0.44<br>(0.151) | 0.57<br>(0.235) | **0.91**<br>**(0.020)** | <0.001 |
| SPC | **1.00**<br>**(0.000)** | 0.98<br>(0.009) | 0.55<br>(0.195) | 0.99<br>(0.004) | 0.99<br>(0.002) | 0.82<br>(0.131) | <0.001 |
| Log loss | 0.236<br>(0.025) | 0.209<br>(0.038) | .. | 0.166<br>(0.048) | **0.152**<br>**(0.053)** | .. | <0.001 |
| F1-score | 0.14<br>(0.050) | 0.52<br>(0.150) | 0.33<br>(0.117) | 0.57<br>(0.178) | **0.64**<br>**(0.237)** | 0.56<br>(0.142) | <0.001 |
| MBS | 0.064<br>(0.008) | .. | .. | **0.046**<br>**(0.015)** | .. | .. | <0.001 |
| AUC | 0.85<br>(0.057) | .. | .. | **0.93**<br>**(0.055)** | .. | .. | <0.001 |
| **UTI** |  |  |  |  |  |  |  |
| PPV | 0.76<br>(0.036) | 0.72<br>(0.027) | 0.60<br>(0.045) | **0.82**<br>**(0.063)** | 0.81<br>(0.066) | 0.73<br>(0.080) | <0.001 |
| SEN | 0.63<br>(0.034) | 0.71<br>(0.070) | **0.90**<br>**(0.016)** | 0.77<br>(0.068) | 0.80<br>(0.058) | **0.90**<br>**(0.013)** | <0.001 |
| SPC | **0.88**<br>**(0.017)** | 0.83<br>(0.019) | 0.62<br>(0.079) | 0.90<br>(0.034) | **0.88**<br>**(0.041)** | 0.79<br>(0.097) | <0.001 |
| Log loss | 0.464<br>(0.029) | 0.454<br>(0.034) | .. | 0.36<br>(0.064) | **0.342**<br>**(0.074)** | .. | <0.001 |
| F1-score | 0.69<br>(0.035) | 0.72<br>(0.043) | 0.72<br>(0.032) | 0.79<br>(0.065) | **0.80**<br>**(0.061)** | **0.80**<br>**(0.056)** | <0.001 |
| MBS | 0.150<br>(0.012) | .. | .. | **0.113**<br>**(0.025)** | .. | .. | <0.001 |
| AUC | 0.86<br>(0.025) | .. | .. | **0.92**<br>**(0.044)** | .. | .. | <0.001 |
| **PSUTI** |  |  |  |  |  |  |  |
| PPV | **1.00**<br>**(0.000)** | 0.74<br>(0.098) | 0.15<br>(0.094) | 0.89<br>(0.134) | 0.78<br>(0.119) | 0.41<br>(0.251) | <0.001 |
| SEN | 0.05<br>(0.019) | 0.45<br>(0.179) | 0.89<br>(0.049) | 0.52<br>(0.167) | 0.69<br>(0.212) | **0.93**<br>**(0.027)** | <0.001 |
| SPC | **1.00**<br>**(0.000)** | 0.99<br>(0.003) | 0.61<br>(0.160) | **1.00**<br>**(0.002)** | 0.99<br>(0.002) | 0.82<br>(0.203) | 0.023 |
| Log loss | 0.142<br>(0.014) | 0.118<br>(0.022) | .. | 0.087<br>(0.029) | **0.079**<br>**(0.033)** | .. | <0.001 |
| F1-score | 0.10<br>(0.035) | 0.54<br>(0.180) | 0.24<br>(0.120) | 0.65<br>(0.181) | **0.73**<br>**(0.183)** | 0.52<br>(0.247) | <0.001 |
| MBS | 0.035<br>(0.003) | .. | .. | **0.022**<br>**(0.008)** | .. | .. | <0.001 |
| AUC | 0.89<br>(0.048) | .. | .. | **0.95**<br>**(0.047)** | .. | .. | <0.001 |
| **Other** |  |  |  |  |  |  |  |
| PPV | 0.81<br>(0.044) | 0.73<br>(0.045) | 0.51<br>(0.042) | **0.82**<br>**(0.077)** | 0.78<br>(0.062) | 0.62<br>(0.085) | <0.001 |
| SEN | 0.35<br>(0.042) | 0.54<br>(0.092) | **0.89**<br>**(0.015)** | 0.62<br>(0.084) | 0.68<br>(0.124) | **0.89**<br>**(0.018)** | <0.001 |
| SPC | **0.96**<br>**(0.007)** | 0.90<br>(0.024) | 0.56<br>(0.094) | 0.93<br>(0.027) | 0.91<br>(0.019) | 0.72<br>(0.127) | 0.011 |
| Log loss | 0.486<br>(0.035) | 0.467<br>(0.045) | .. | 0.396<br>(0.070) | **0.372**<br>**(0.082)** | .. | <0.001 |
| F1-score | 0.49<br>(0.049) | 0.61<br>(0.068) | 0.65<br>(0.038) | 0.70<br>(0.083) | **0.73**<br>**(0.097)** | **0.73**<br>**(0.065)** | <0.001 |
| MBS | 0.156<br>(0.014) | .. | .. | **0.123**<br>**(0.026)** | .. | .. | <0.001 |
| AUC | 0.83<br>(0.039) | .. | .. | **0.89**<br>**(0.058)** | .. | .. | <0.001 |

Figure 4a illustrates the receiver operating characteristics (ROC) curves of the XGBoost model. The XGBoost model was the best performing model based on AUC, multi class Brier Score, Log loss, and F1-score. The model generally shows strong performance in predicting the patients’ site of infection, with the highest AUC observed for predicting airway infections and urine contamination, while the lowest AUC is seen in the “Other infections” class. The class that achieves the best balance between sensitivity and specificity is the Urine infections as this class has the highest F1-score. Figure 4b-d depicts the confusion matrices for the XGBoost model in its naïve, VA calibrated, and CRC-VA calibrated states. It is evident that VA calibration substantially increases the number of true positives (TP). Controlling the sensitivity to at least 90% on average using CRC results in an increased number of TPs, but also increases the prediction set size, as indicated by the elevated number of False Positives (FP).

**Figure 3.**
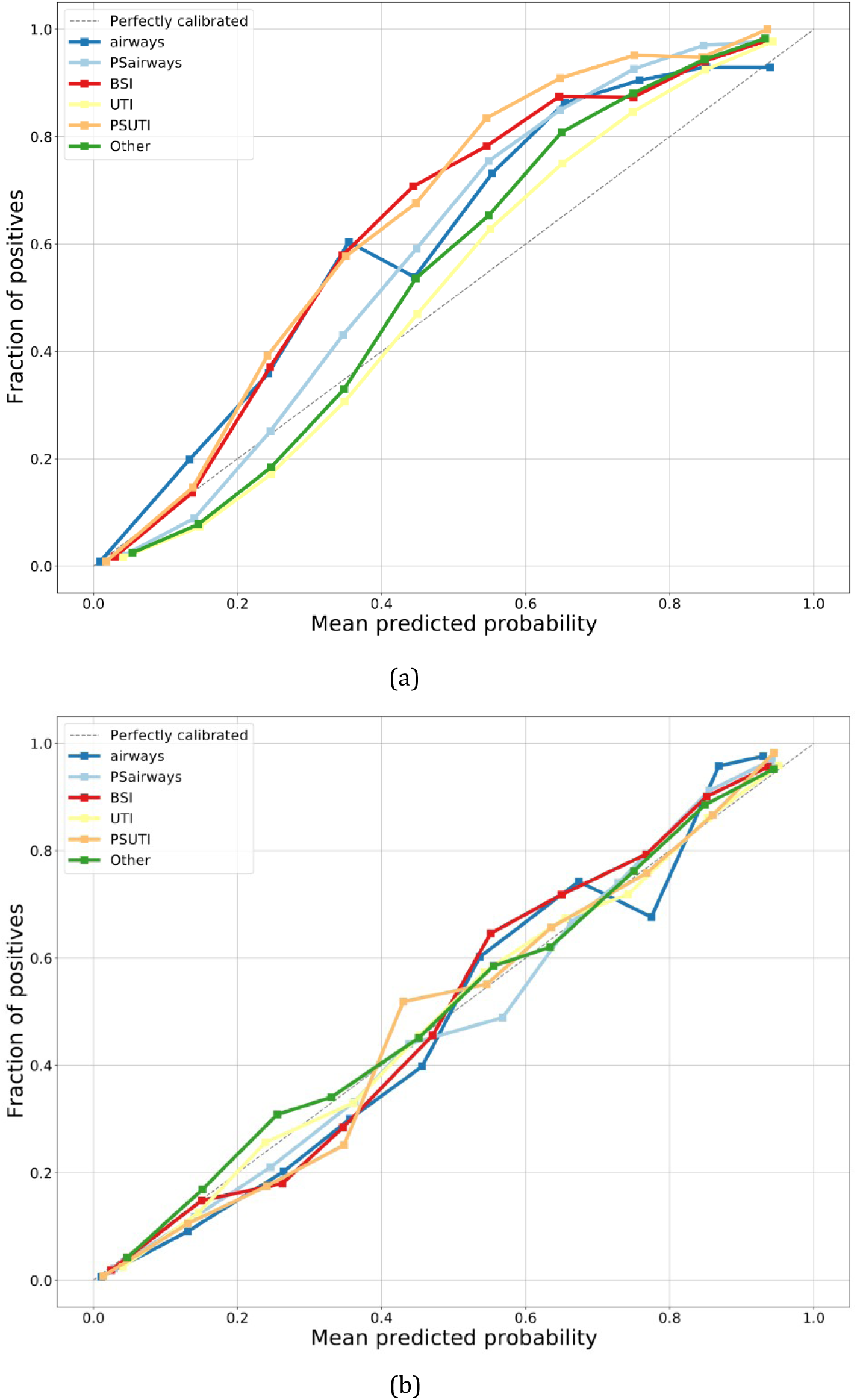
(a) Naïve XGBoost (b) VA calibrated XGBoost. Micro averaged calibration curves for the XGBoost model, showing the mean predicted probability vs. the fraction of positives. (a) significantly underestimates the infection probability and is therefore untrustworthy, while (b) is well calibrated and therefore provides trustworthy probability estimates. The blue lines represent airway infections (airways) and common contaminants of the airways (PSairways). The red line represents blood stream infections (BSI). The yellow lines represent urinary tract infections (UTI) and common contaminants of the urinary tract (PSUTI). The green line represents other types of infection (Other).

**Figure 4.**
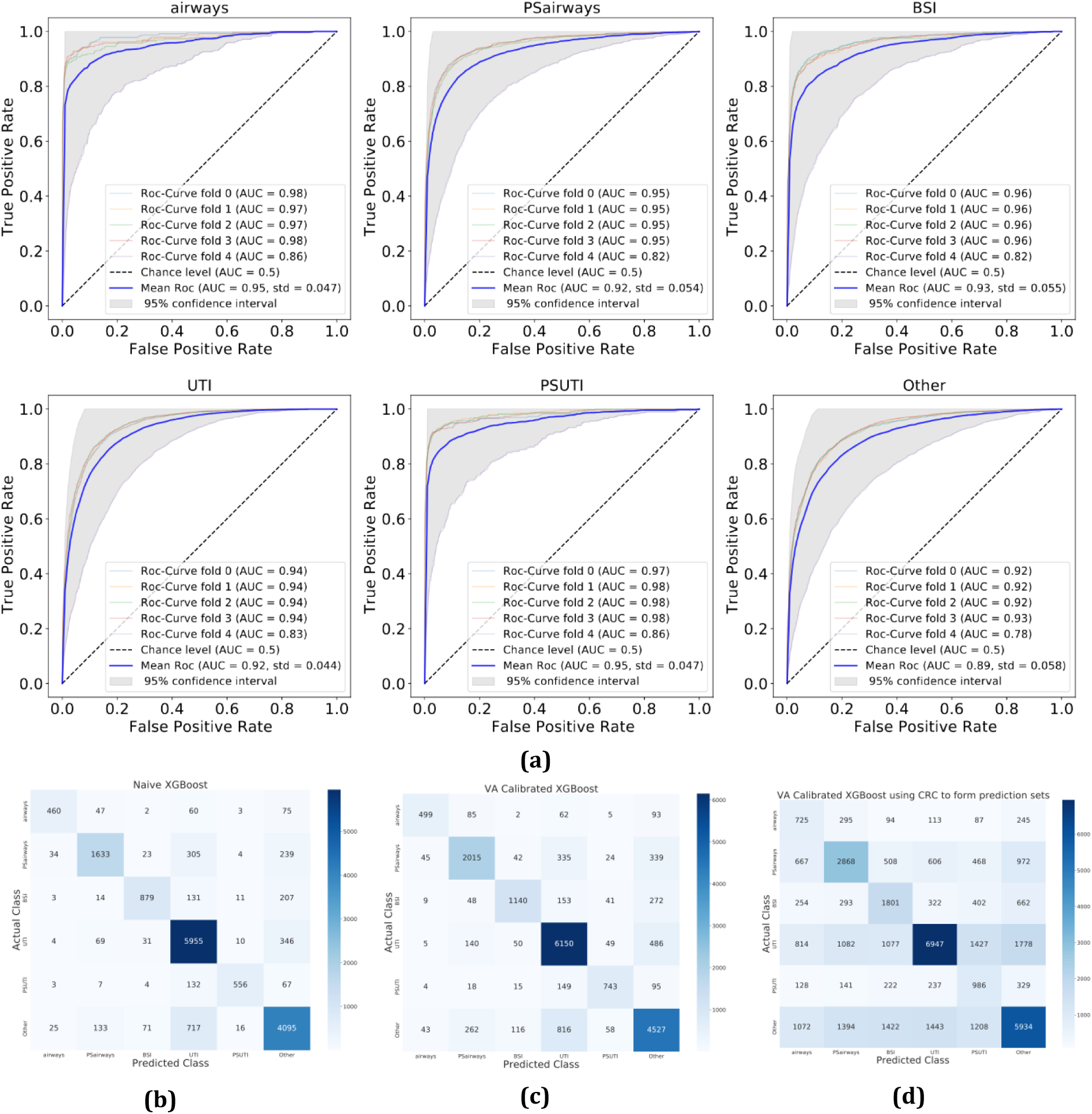
Overview of the XGBoost model performance on the test sets. (a) receiver operating characteristics (ROC) for each infection class using 5-fold cross validation. (b), (c), (d) Confusion matrices for the XGBoost model in its naïve, VA calibrated, and CRC-VA calibrated form respectively. The confusion matrices are summed across the 5-fold cross validation (for reference a theoretically perfect confusion matrix can be seen in S8) From the confusion matrices it is clear that both calibration and CRC uncertainty based prediction sets increase the number of correct predictions. VA: Venn-ABERS, CRC: conformal risk control, Airways: airway infections, BSI: blood stream infection, UTI: urine tract infection, PS: organisms that are usually not pathogens and common contaminants, AUC: area under the ROC curve.

The CRC method mathematically signals uncertainty in a manner similar to human intuition. Figure 5 displays the prediction set sizes at different levels of *α*. When *α* = 0.01, corresponding to a sensitivity of at least 99% on average, the model includes a larger proportion of the label space in the prediction set. Higher levels of *α* results in smaller prediction sets with a larger risk of missing the true class. At *α* = 0.3 corresponding to a sensitivity of at least 70% on average, the median prediction set size is one. It is also evident that the CRC method can produce empty prediction sets when the predicted probability of each class is below the probability threshold, indicating lack of confidence in predicting a class label.

**Figure 5.**
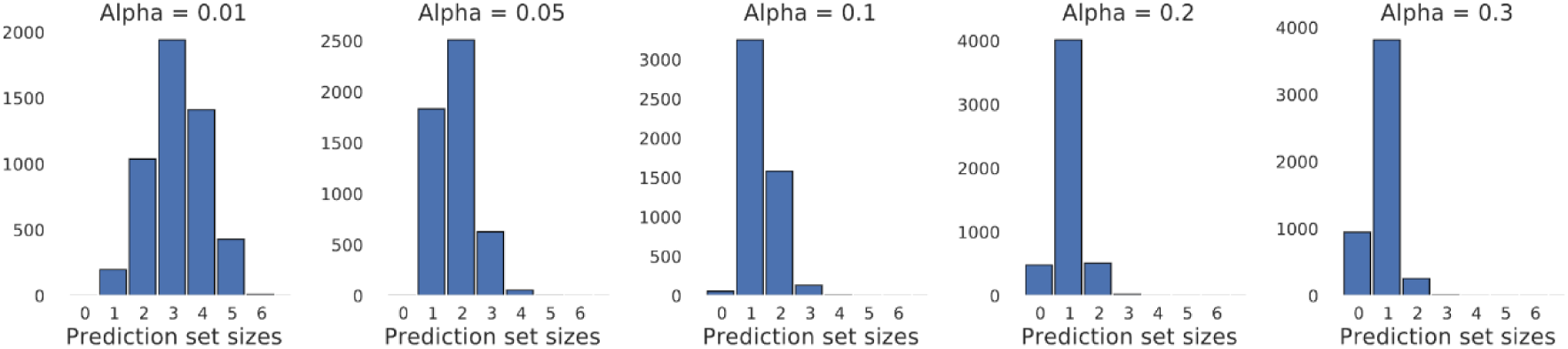
Prediction set sizes at different levels of α when applying the conformal risk control method. α corresponds to the user specified error tolerance, from left to right the figure shows prediction set sizes at 1, 5, 10, 20 and 30% error rates. Here It can be seen that large user tolerated error rates results in bolder or overconfident prediction sets, while small user tolerated error rates result in large prediction sets.

Active learning (AL is a framework ^19^ aimed at maximising model performance while minimizing the number of annotated samples required. Based on the available dataset, we estimate the amount of data needed to achieve the expected results using a pool based AL strategy. Figure 6 shows that the XGBoost model requires approximately 2,000 positive samples per class to approach the maximal obtainable sensitivity. This can also be seen in figure S 14 which depicts the relationship between the std. of micro averaged performance metrics from the XGBoost model.

**Figure 6.**
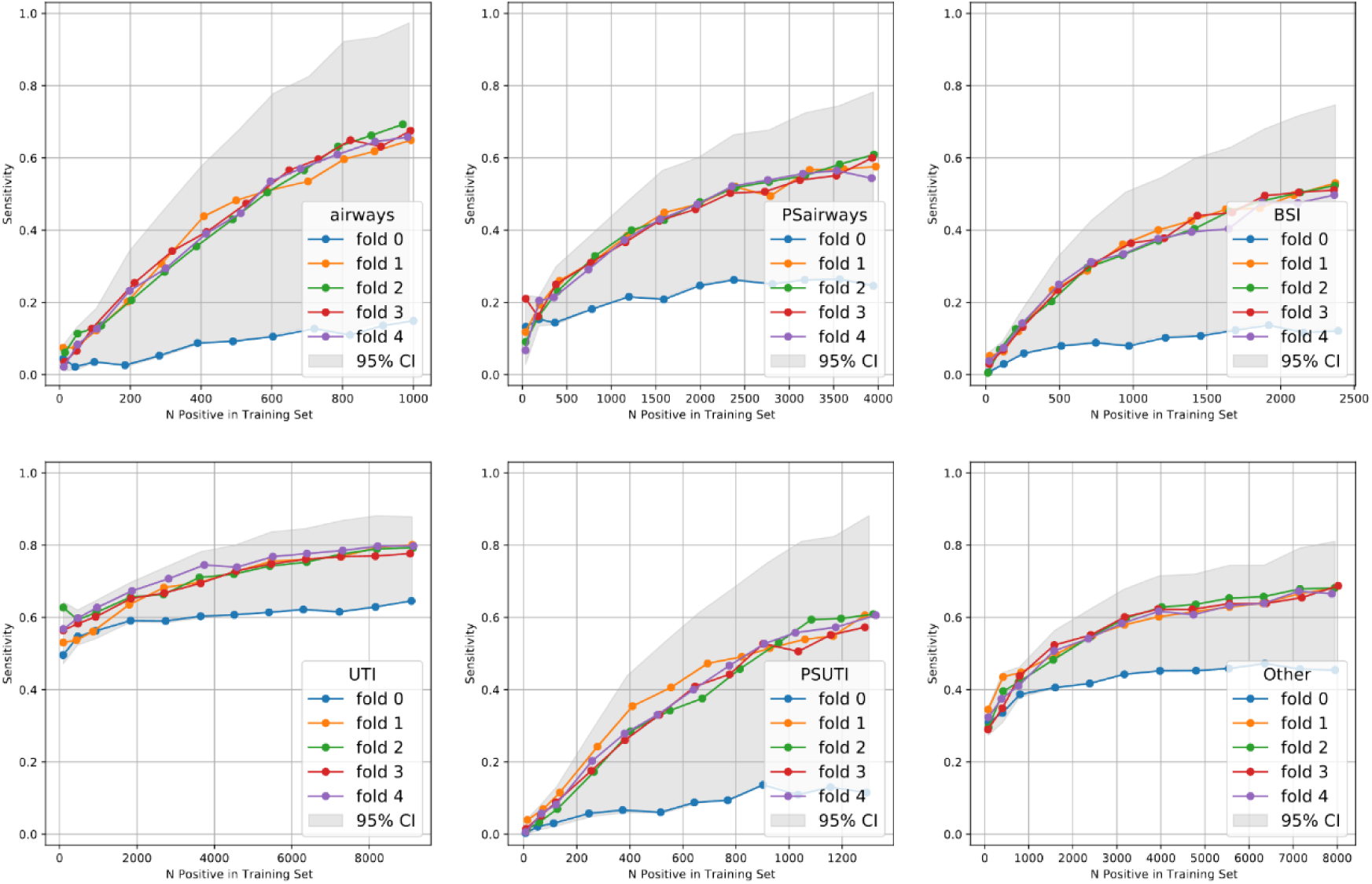
Sensitivity of the Naïve XGBoost model plotted vs. the number of positives in the training dataset for each infection class, when the model is trained with different training dataset sizes. Here it is seen that model performance starts to stagnate at approximately 1,000-2,000 positive observations. Airways is airway infections, BSI is blood stream infections, UTI are urine tract infections, PS denotes organisms that are usually not pathogens and common contaminants,, and the “Other” class is an ensemble of other types of infection Similar results for other performance metrics can be seen in S7-S13.

The ablation study, removing hematopoietic patients and patients with low leucocyte count yielded similar results based on the performance metrics, only varying AUC by a few percentage points (absolute difference in AUC was 1.17 ± 1.17 pp. ± std.).

The SHAP-plots in Figure 7 illustrate feature importance across the different classes of infection. Figure 7a ranks features by their SHAP-value, highlighting their importance. It is evident that the model heavily relies on information about the patient’s ability to breathe and oxygenate their blood, as O2 therapy and blood oxygen saturation are amongst the most important features.

**Figure 7.**
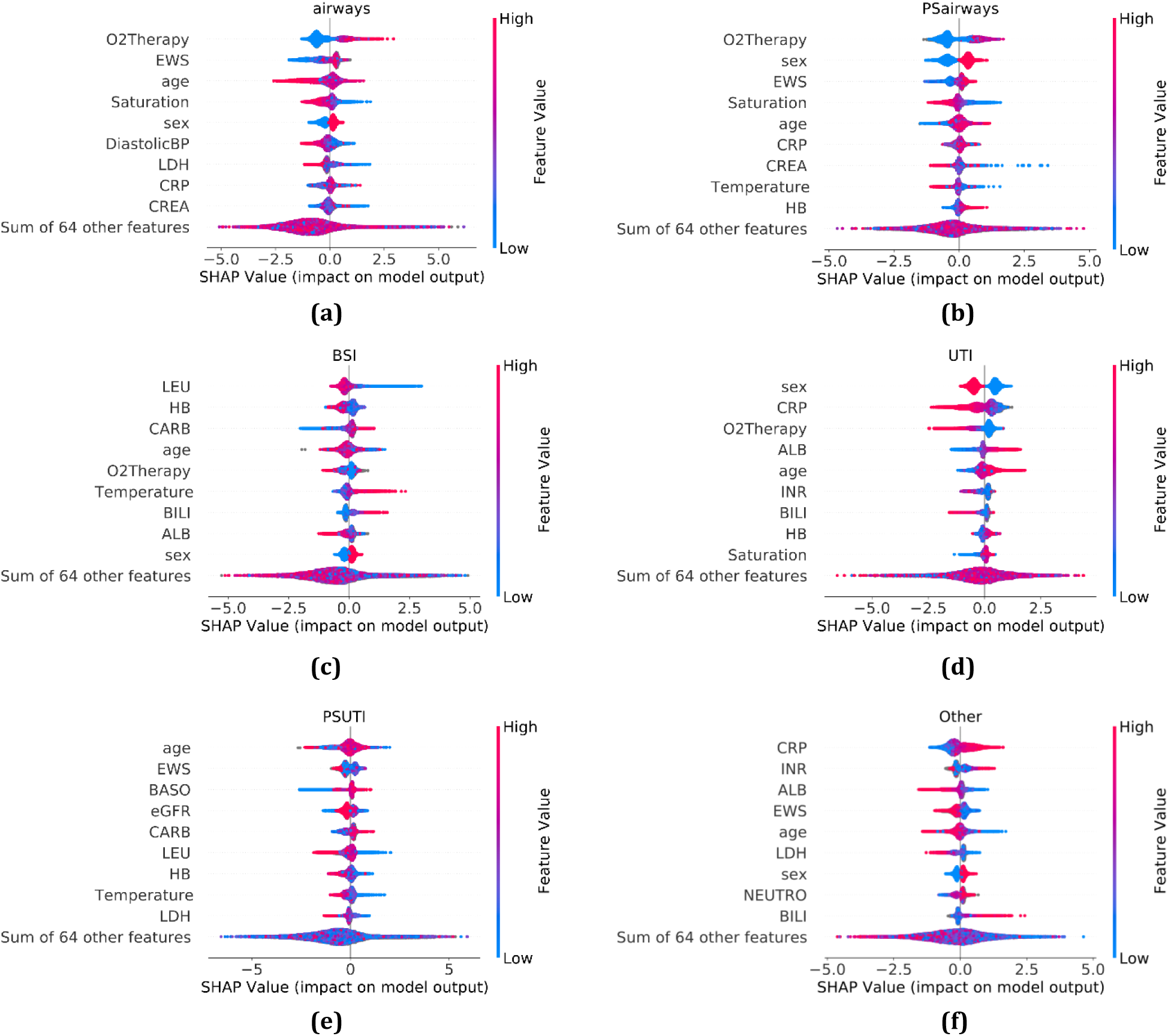
SHAP summary plots displaying feature importance of the 9 most important features as determined by SHAP-value. The SHAP-value is equivalent to the log-odds of predicting the class. Here it can be seen that the model generally highlights known risk factors associated with each infection focus. Sex is encoded categorically as, male=1, female=0. airways: airway infection, BSI: blood stream infection, UTI: urine tract infection, PS: oranisms that are usually not pathogens and common contaminants, O2Therapy: O2 Therapy, EWS: Early warning score, age: patient age in years, Saturation: blood O2 saturation percentage, LDH: lactate dehydrogenase, CREA: creatinine, eGFR: estimated glomerular filtration rate, DiastolicBP: diatolic blood pressure, LEU: leukocyte count, LYMFO: lymphocyte count, Temperature: patient body temperature, CARB: Carbamide, BILI: bilirubin, ALB: albumin, HB: hemoglobin, CRP: C-reactive protein, HR: Heart rate, SystolicBP: Systolic blood pressure, INR: International normalized ratio blood test, BASO: Basophilocyte count, GLU: Glucose, NEUTRO: Neutrophil count.

Figure 7c depicts the feature importance related to BSIs. The two most important features for predicting BSIs are the leukocyte (LEU) count and the hemoglobin concentration (HB). Specifically, a low LEU count corresponds to a high risk of BSI compared to all other classes of infection. While bacterial infections typically cause elevated LEU counts ^22^, this result may be explained by the large group of immunocompromised patients at Rigshospitalet. Furthermore, the patients’ temperature appears to be a predictor of BSI. This aligns with the literature, as patients with BSI are known to have elevated body temperatures, except in cases where the blood circulation is affected, which can cause hypothermia ^23^.

Figure 7d shows that the greatest risk factor when predicting UTI is the patient sex, with female encoded as zero and male as one. Furthermore, high levels of C-reactive protein (CRP) correspond to a lower chance of UTI compared to the other classes of infection, while a high concentration of albumin (ALB) indicates a higher likelihood of predicting UTI.

## 4. Discussion

The aim of this study was to develop a data-driven predictive probabilistic model that can learn from historic real patient data and predict the focus of infection within a reasonable margin of error and account for how uncertainty in the data translate to predictive uncertainty to improve model reliability. The best performing ML-model was the XGBoost model, achieving a Log loss of 0.219 and an AUC of 0.93, which is infrequently observed within the field of bacterial infections and indicative of good model performance. This model also showed a decent balance between sensitivity and specificity as seen by the F1-score of 0.71, meaning that the model performance is acceptable, both when measured on how well it predicts the correct label and how well it excludes the incorrect labels. Metrics such as AUC, F1-score, etc. that are based on classification outcomes are however known to be misleading^24^. For example, a model can achieve high AUC while still producing poorly calibrated or overconfident probability estimates and can therefore not be used as sole indicators of performance. In contrast, metrics like Log loss and Brier score directly considers the predicted probabilities and calibration hereof. Log loss penalizes incorrect and overconfident predictions more heavily, while Brier score measure the mean squared difference between predicted probabilities and actual outcomes across all classes. These metrics are therefore more appropriate when the aim is to produce reliable, real-world probabilistic predictions.

The XGBoost model also outperforms the SVM when compared on Log loss and Brier score. The naïve model state did, however, prove to be somewhat badly calibrated, nevertheless using Venn-ABERS (VA) calibration a reasonably well calibrated model with a multi class brier score of 0.366 and a Log loss of 0.205 was obtained. Applying conformal risk control (CRC) in this setting guarantees a sensitivity of at least 90% on average, meaning that in nine out of ten cases, the correct prediction is included in the prediction set, while also not creating excessively large prediction sets. Compared to similar studies in the field of bacterial infections VA-calibration and CRC results in a considerably improved model performance.

Furthermore, it was found, through active learning that studies should include a minimum of 1,000 positive observations pr. class and preferably 2,000 observations to achieve satisfactory model quality and robust uncertainty quantification. A dataset of this magnitude may be difficult to obtain in most hospital settings. However, considering the active learning results, this dataset size is essential for training a robust ML-model using XGBoost, when applied to patient data with the purpose of predicting bacterial infection focus.

Adopting ML to clinical practices requires a high level of trustworthy artificial intelligence^25^ (AI). In a clinical setting clinicians and diagnosticians are responsible for human lives. For the clinicians and diagnosticians to trust the predictions made by AI and ML-models, the models need to be built with transparency, robustness, and a high level of explainability ^25^. To achieve this one should address known biases in the dataset, potential strengths, and weaknesses of the ML-models, and provide explanations for the model outputs.

The dataset in this study comes with a handful of natural biases. The data is recorded at Rigshospitalet in Denmark. Rigshospitalet is a highly specialised tertiary hospital, patients therefore often present with numerous comorbidities.

This makes diagnostics increasingly difficult, but it also introduces biases towards sick people in the dataset. The input features were transformed to resemble the gaussian distribution, since this is a common choice for input features. However, we acknowledge that an ablation study assessing the impact of this transformation would provide further insight. The ML-labels used in this study could also introduce biases. The labels are generated using the sample origin and the bacterial species. However, this combination is likely not perfectly true to reality and could potentially introduce a level of epistemic errors into the dataset. To increase trustworthiness and diagnostic value of the ML-models the labels should be redefined using other more strict definitions of contamination.

The findings align well with previous knowledge, expected findings, and demonstrate that the ML-modelling can support identifying clinically relevant factors for predicting the different foci for bacterial infections. The ML-model’s heavy reliance on O2 therapy and blood oxygen saturation for predicting respiratory infections, the identification of leukocyte counts and hemoglobin concentration as important features for BSI prediction, and the identification of patient sex as the most important factor for UTI prediction is clinically sensible and consistent with epidemiological data. The inverse relationship between CRP levels and UTI prediction, as well as the positive association with albumin levels, may reflect the model’s ability to distinguish UTIs from more severe systemic infections. This nuanced interpretation of biomarkers demonstrates the ML-model’s sophistication in differentiating between infection types. These results are promising since the active learning framework suggests that approximately 2,000 positive samples per class are needed to achieve near-optimal model performance. This information is valuable for future studies, as it provides a clear guideline for dataset size requirements. It also indicates that the current study likely had sufficient data to develop a robust model.

The naïve models proved to be badly calibrated. This issue was handled using VA-calibration. Badly calibrated models could potentially decrease the trustworthiness of the model and ML in general, as users empirically will start to doubt the ML-models if it consistently outputs either lower or higher chance of bacterial infection than what the user observes.

The usage of tools in this study are chosen with clinical application in mind. Specifically, the usage of conformal predictions to generate statistically robust prediction sets is of potentially high value in a clinical setting. Many ML-methods do not provide uncertainty estimates for their predictions. Applying the conformal risk control (CRC) method generates prediction sets of varying size as seen in Figure 5. This is clinically useful as the framework not only provides statistically sound uncertainty guaranties for the user specified performance metrics, but also allows the ML-models to generate prediction sets of varying size, which essentially signals uncertainty in a human like fashion and is known to significantly improve accuracy in human-in-the-loop systems^26^, where humans make decisions based on expert knowledge and ML-outputs.

Prediction sets can be a great way of signalling uncertainty, but the size of the prediction sets needs to be discussed. For example, using CRC it is possible to guarantee a certain level of sensitivity. Controlling the sensitivity to 99% for the XGBoost model results, by definition, in few predictions sets where the correct label is excluded. This is however achieved by outputting half the label space for median observations. In practice large prediction sets do not provide any direct diagnostic value to the clinicians or diagnosticians, it does however indicate high uncertainty, which warrants further examination of the patient. Reducing the sensitivity to 70% mostly resulted in prediction set sizes of two or less. This size is potentially more useful as it significantly narrows the potential search field. The issue with this setting is that the model essentially guarantees an error rate of approximately 30%. When using CRC in practice it is therefore important to find the right balance between prediction set size and error rate, such that the prediction sets are narrow enough that clinicians and diagnosticians can extract actual value from them, but still wide enough to provide a reasonably low error rate. Initially CRC might not seem like a powerful tool solely based on performance metrics, however considering that CRC provides any model with statistically robust uncertainty guaranties on the sole condition that new observations stem from the same distribution as the one provided by the calibration dataset, it should perhaps not only be considered a powerful tool, but a necessity when applying ML in a health care setting.

In terms of explainability the XGBoost model has been analysed using SHAP-plots. SHAP-plots are widely used as a tool for explainable AI. SHAP-analysis assumes feature independence, this is almost never achievable in real life applications, and the SHAP-analysis should therefore be interpreted with a pinch of scepticism^21^. From the SHAP-analysis it is also clear that the dataset contains human introduced biases, as O2 therapy is deemed as the most important feature when predicting airway infections. This feature is controlled by the diagnosticians and is therefore naturally biased towards the human decision maker. Nevertheless, the majority of the features deemed important by the model are expected and known biomarkers of bacterial infection (age^27^, LDH^28^, LEU^22^, temperature^23^, sex, and CRP).

Our study has several limitations. It is a retrospective design, which may limit the ability to control for confounding factors, and the model was not validated on an external dataset from a different hospital or time period, which would have provided stronger evidence of its generalizability. The low prevalence of positive blood cultures in the dataset may affect the model’s performance, particularly its positive predictive value. Additionally, the model does not incorporate important clinical information such as comorbidities or presence of indwelling devices, which are known risk factors for bloodstream infections.

However, this study combines advanced machine learning techniques (XGBoost) with conformal prediction methods (VA calibration and CRC) to create a more reliable and interpretable model.

The use of CRC to generate prediction sets of varying sizes based on uncertainty is an approach developed for, inter alia, clinical decision support, mimicking human-like signalling of uncertainty and the comprehensive analysis of bacterial infections using a wide range of biochemical and vital sign data is more extensive than previous studies in this field.

The integration of trustworthy AI principles, addressing model uncertainty and providing explainable results, represents a significant step towards clinical application of ML in infection diagnosis. Thus, this study provides valuable insights into the data requirements for developing similar models in the future, which is crucial for advancing the field of ML in clinical microbiology.

## 5. Conclusion

The XGBoost model achieved high performance (F1-score: 0.71, AUC: 0.93, Log loss: 0.219, MBS: 0.389) in predicting the focus of bacterial infections using electronic health record data and biochemical markers. Venn-ABERS calibration and conformal risk control significantly improved model calibration and guaranteed a sensitivity of at least 90% on average. We estimate that approximately 2,000 positive samples per infection class are needed to achieve optimal model performance, as determined by active learning analysis. The model identified clinically relevant predictors for different infection types, such as oxygen therapy for respiratory infections and leukocyte count for bloodstream infections, which aligns well with clinical observations. Conformal prediction methods can generate variable-size predictions sets, signalling uncertainty in a human-like fashion and potentially improve clinical decision-making. Despite limitations such as its retrospective design and lack of external validation, this datadriven ML-approach of combining classical ML-models like XGBoost with conformal prediction methods provides valuable insights into data requirements and methodological approaches for developing trustworthy AI systems in clinical microbiology. These findings may have significant implications for improving antibiotic stewardship and patient outcomes in the management of bacterial infections.

## Author contribution

Conceptualization: Jacob Bahnsen Schmidt, Frederik Boëtius Hertz, Karen Leth Nielsen, Allan Peter Engsig-Karup

Data curation: Jacob Bahnsen Schmidt, Dmytro Strunin, Nikolai Søren Kirkby, Steen Christian Rasmussen

Formal analysis: Jacob Bahnsen Schmidt Investigation: Jacob Bahnsen Schmidt

Methodology: Jacob Bahnsen Schmidt, Frederik Boëtius Hertz, Karen Leth Nielsen, Allan Peter Engsig-Karup

Ethical approval: Frederik Boëtius Hertz.

Resources: Frederik Boëtius Hertz, Karen Leth Nielsen Software: Jacob Bahnsen Schmidt, Dmytro Strunin Formal analysis: Jacob Bahnsen Schmidt

Supervision: Frederik Boëtius Hertz, Karen Leth Nielsen, Allan Peter Engsig-Karup

Validation: Jacob Bahnsen Schmidt, Dmytro Strunin, Jesper Qvist Thomassen, Steen Christian Rasmussen

Visualization: Jacob Bahnsen Schmidt

Project administration: Karen Leth Nielsen, Frederik Boëtius Hertz, Allan Peter Engsig-Karup Writing – Original Draft Preparation: Jacob Bahnsen Schmidt, Frederik Boëtius Hertz.

Writing – Review & Editing: Jacob Bahnsen Schmidt, Frederik Boëtius Hertz, Karen Leth Nielsen, Dmytro Strunin, Nikolai Søren Kirkby, Jesper Qvist Thomassen, Steen Christian Rasmussen, Ruth Frikke-Schmidt, Allan Peter Engsig-Karup

Materials & Correspondence: Jacob Bahnsen Schmidt

## Ethical approval

There were no ethical concerns for this study. This was an observational, non-intervention study. The extraction of data from registries was approved by the local data protection agency (Journal-nr.: R-21015888) and registered in Pactius (P-2020-743).

## Data Sharing

All relevant data are within the manuscript and its Supporting Information files. We are unable to share sensitive data instances due to compliance with the General Data Protection Regulation (GDPR) and Danish legislation. These regulations mandate that the privacy and confidentiality of individuals must be protected, and sensitive data should not be made publicly available without appropriate safeguards. Therefore, we cannot provide access to the sensitive data instances used in this study to ensure compliance with these regulations.

## Code Availability

Access to the code used for the statistical design and ML-process can be made available on reasonable request to the authors.

