## Supplementary for "Conformal Prediction and Venn-ABERS Calibration for Reliable Machine Learning-Based Prediction of Bacterial Infection Focus"

**S1**

*S1 Table showing the cut-off values for outlier removal, when filtering the biochemical variables and the vital parameters. Empty fields mean that there are no relevant limits for that feature.*

| Variable Code | Variable Name | Lower boundary | Upper boundary | Note |
|---|---|---|---|---|
| ABGLU | Glucose Arterial blood | | | |
| ABPH | Arterial blood pH | | | |
| ALAT | Alanine aminotransferase | | 92 | |
| ALB | Albumin | | | |
| AMYL | Amylase | | 198 | |
| APTT | Activate partial thromboplastin time | | 100 | |
| ASAT | Aspartate transaminase | | 500 | |
| BASO | basophilocyte count | | 5 | |
| BILI | Bilirubin | | | |
| BILIKON | Conjugated bilirubin | | | |
| BILIUK | Unconjugated bilirubin | | | |
| CA | Calcium | | | |
| CAI | Ionized calcium | | | |
| CARB | Carbamide | | | |
| CHOL | Cholesterol | | | |
| CL | Chloride | | | |
| CREA | Creatinine | | | |
| CRP | C-reactive protein | | | |
| DIMER | Fibrin D-Dimer | | | |
| ECVBE | Base excess Ecv | | | |
| EOS | Eosinophil count | | | |
| ERY | Erythrocyte count | | | |
| FERRITIN | Ferritin | | 2500 | |
| GGT | Gamma-Glutamyl Transferase | | | |

| Variable Code | Variable Name | Lower boundary | Upper boundary | Note |
|---|---|---|---|---|
| GLU | Glucose | 0.001 | | |
| HAPTO | Haptoglobin | | | |
| HB | Hemoglobin | 0 | | |
| HBA1C | Glycated Hemoglobin | | | |
| HCO3 | Hydrogencarbonat | | | |
| HDL | High Density Lipoprotein cholesterol | | | |
| INR | International normalized ratio blood test | | | |
| JERN | Iron | 0.001 | | |
| K | Potassium | | | |
| KBGLU | Capillary blood glucose | | | |
| LDH | Lactate dehydrogenase | | 2500 | |
| LDL | Low Density Lipoprotein cholesterol | | | |
| LEU | Leukocyte count | | | |
| LYMFO | Lymphocyte count | | 25 | |
| MCH | Mean corpuscular hemoglobin | | | |
| MCHC | Mean corpuscular hemoglobin concentration | | | |
| MG | Magnesium | | | |
| MONO | Monocyte count | | | |
| NA | Sodium | | | |
| NEUTRO | Neutrophil count | | | |
| PHOS | Phosphate | | | |
| PROCAL | Procalcitonin | | 25 | |
| TRANSJBG | Transferrin saturation | | | |
| TRIG | Triglyceride | | | |
| TSH | Thyroid Stimulating Hormone | | | |
| eGFR | estimated Glomerular Filtration Rate | | | |
| BMI | Body Mass Index | 10 | 75 | |
| Diastolic BP | Diastolic Blood Pressure | 0 | | |
| HR | Heart Rate | 0 | 350 | |

| Variable Code | Variable Name | Lower boundary | Upper boundary | Note |
|---|---|---|---|---|
| O2Therapy | Oxygen therapy | 0 | 150 | |
| RespFreq | Respiration frequency | 0 | 100 | |
| Saturation | Blood oxygen saturation | 0 | 100 | |
| Systolic BP | Systolic Blood Pressure | 0 | 400 | |
| Temperature | Patient Temperature | 10 | 42.3 | |
| Weight | Weight | 20 | | If age is > 15 |
| Weight | Weight | 25 | | If age is > 20 |
| Weight | Weight | 0.2 | 250 | |

**S2**

The study contains three engineered features, BAS, ABAS and MBAS. These three features have previously proven to have a positive impact on the overall predictive ability of similar models, as shown by Marandi et al. in their study on BSI's.[27]

BAS stands for biochemical abnormality score and is used to describe the patients' level of abnormality related to the biochemical variables. Equation 2 shows the formula used to calculate BAS, where the normal ranges that are referred to are the medically considered normal ranges at Rigshospitalet, defined by Labportalen at https://labportal.rh.dk/Labportal.asp?ShowStart=Y. A full table with the biochemical normal ranges can be seen in S5.

$$\text{BAS} = \frac{\text{Number of biochemical variables outside the normal range}}{\text{Number of biochemical variables}} \cdot 100 \qquad \text{Equation 2}$$

ABAS is the adjusted biochemical abnormality score calculated as in Equation 3, where the abnormality level is adjusted such that it only accounts for the measured variables.

$$A\text{BAS} = \frac{\text{Number of biochemical variables outside the normal range}}{\text{Number of available biochemical variables}} \cdot 100 \qquad \text{Equation 3}$$

MBAS is the minimal biochemical abnormality score, describing the level of abnormality given a specific selected subset of commonly used biochemical variables and is calculated as in Equation 4.

$$\text{MBAS} = \frac{\text{Number of selected biochemical variables outside the normal range}}{\text{Number of selected biochemical variables}} \cdot 100 \qquad \text{Equation 4}$$

Where the selected biochemical variables refer to a list of five commonly used biochemical variables:

- **ALAT**, Alanine transaminase is a biomarker for acute liver damage.
- **CRP**, C-reactive protein is an acute phase protein produced during inflammation.
- **CREA**, Creatinine is a biomarker for kidney function.
- **LEU**, Number of Leukocytes (white blood cells).
- **PROCAL**, Procalcitonin is a potential biomarker for bacterial infections.

**S3**

SVM's aim to solve classification tasks by constructing hyperplanes in high or infinite dimensional spaces. The SVM cannot handle datasets with missingness. The dataset is therefore further pre-processed imputating missingness using the feature median. Furthermore, the SVM performance relies on the input features to have similar variance. The dataset is therefore scaled using a standard scaler, such that each feature has zero mean and unit variance. The SVM was optimised using a narrow grid search. The optimal kernel function was the Radial Basis Function (RBF) kernel. The parameters C and $\gamma$ was set to 1 and $\frac{1}{\rho Var(X)}$ respectively, where $\rho$ is the number of features.

**S4**

In practice, it is desirable to produce a model that can provide uncertainty guaranties. This study has therefore used the Conformal Risk Control (CRC) procedure from the conformal prediction framework.[17] CRC is a method that utilises conformal predictions in a multi-label setting, in order to control a risk function. Mathematically, this is formulated as in Equation 5.

$$\mathbb{E}[l(\mathcal{C}(X_{n+1}), Y_{n+1})] \leq \alpha$$

*Equation 5*

Where $\alpha$ is a user specified error rate, $l(\mathcal{C}(X_{n+1}), Y_{n+1})$ is a bounded loss function, that decreases when the size of the prediction sets $\mathcal{C}(X_{n+1})$ increases. $Y_{n+1}$ is the ground truth label set of a new observation. In the multi-label setting the sets will be defined as a subset of K classes $Y_i \subseteq \{1, \ldots, K\}$. The goal is then to produce a multi-label classifier $f : \mathcal{X} \rightarrow [0,1]^K$ such that the prediction sets contain a sizeable proportion of true labels.

This is achievable by post processing model outputs such that a probability threshold is obtained for each class, $C_\lambda(x) = \{k : f(X)_k \geq 1 - \lambda_k\}$, such that only classes with adequately high scores are included in the prediction sets. This study used CRC to control the sensitivity of the ML-models, utilising the loss-function seen in Equation 6.

$$l(\mathcal{C}_{\hat{\lambda}}(X_{n+1}), Y_{n+1}) = 1 - \frac{|Y_{n+1} \cap \mathcal{C}_{\lambda}(X_{n+1})|}{|Y_{n+1}|} \quad \textit{Equation 6}$$

This study set $\alpha$ = 0.1, which guaranties that the sensitivity is at least 90% on average.

## S5

*S5 Table showing the normal ranges for the biochemical variables as defined by Rigshospitalets labportal available at https://labportal.rh.dk/Labportal.asp?ShowStart=Y.*

| Variable | Sex | Age (years) | Lower boundary | Upper boundary | Unit |
|---|---|---|---|---|---|
| ABGLU | | | 4.2 | 7.2 | mmol/L |
| ABPH | | | 7.37 | 7.45 | pH |
| ALAT | | 0-4 | 5 | 45 | U/L |
| ALAT | Women | 5-17 | 8 | 32 | U/L |
| ALAT | Men | 5-8 | 8 | 27 | U/L |
| ALAT | Men | 9-13 | 8 | 37 | U/L |
| ALAT | Men | 14-7 | 8 | 47 | U/L |
| ALAT | Women | 18-125 | 10 | 45 | U/L |
| ALAT | Men | 18-128 | 10 | 70 | U/L |
| ALB | | 0 | 26 | 34 | g/L |
| ALB | | 1-3 | 34 | 42 | g/L |
| ALB | | 4 | 36 | 48 | g/L |
| ALB | Women | 5-14 | 39 | 47 | g/L |
| ALB | Women | 15-17 | 35 | 47 | g/L |
| ALB | Men | 5-17 | 39 | 50 | g/L |
| ALB | | 18-39 | 36 | 48 | g/L |
| ALB | | 40-69 | 36 | 48 | g/L |
| ALB | | 70-125 | 34 | 45 | g/L |
| AMYL | | 0-1 | | 80 | U/L |
| AMYL | | 2-17 | | 105 | U/L |
| AMYL | | 18-125 | 25 | 120 | U/L |
| APTT | | 0 | 25 | 35 | s |
| APTT | | 1-125 | 25 | 37 | s |
| ASAT | | 0 | 15 | 65 | U/L |
| ASAT | | 1-4 | 10 | 60 | U/L |
| ASAT | | 5-17 | 17 | 46 | U/L |
| ASAT | Women | 18-125 | 15 | 35 | U/L |
| ASAT | Men | 18-125 | 15 | 45 | U/L |
| BASO | | | 0.01 | 0.1 | $10^9$/L |
| BILI | | 0-4 | 5 | 25 | µmol/L |

| Variable | Sex | Age (years) | Lower boundary | Upper boundary | Unit |
|---|---|---|---|---|---|
| BILI | Women | 5-17 | 3 | 18 | µmol/L |
| BILI | Men | 5-13 | 3 | 20 | µmol/L |
| BILI | Men | 14-17 | 3 | 25 | µmol/L |
| BILI | | 18-125 | 5 | 25 | µmol/L |
| BILIKON | | | | 4 | µmol/L |
| BILIUK | Women | | | 17 | µmol/L |
| BILIUK | men | | | 22 | µmol/L |
| CA | | 0 | 2.1 | 2.62 | mmol/L |
| CA | | 1-4 | 2.17 | 2.66 | mmol/L |
| CA | women | 5-13 | 2.26 | 2.58 | mmol/L |
| CA | women | 14-17 | 1.95 | 2.58 | mmol/L |
| CA | men | 5-13 | 2.22 | 2.58 | mmol/L |
| CA | men | 14-17 | 2.1 | 2.58 | mmol/L |
| CA | | 18-125 | 2.15 | 2.51 | mmol/L |
| CAI | | | 1.18 | 1.32 | mmol/L |
| CARB | | 0-1 | 1.8 | 5.4 | mmol/L |
| CARB | | 2-17 | 2.5 | 7.5 | mmol/L |
| CARB | women | 18-49 | 2.6 | 6.4 | mmol/L |
| CARB | women | 50-125 | 3.1 | 7.9 | mmol/L |
| CARB | men | 18-49 | 3.2 | 8.1 | mmol/L |
| CARB | men | 50-125 | 3.5 | 8.1 | mmol/L |
| CHOL | | | | 5 | mmol/L |
| CL | | 0-2 | 91 | 115 | mmol/L |
| CL | | 3-17 | 97 | 108 | mmol/L |
| CL | | 18-125 | 98 | 106 | mmol/L |
| CREA | | 0 | 14 | 34 | µmol/L |
| CREA | | 1-2 | 15 | 31 | µmol/L |
| CREA | | 3-4 | 23 | 37 | µmol/L |
| CREA | women | 5-8 | 28 | 50 | µmol/L |
| CREA | women | 9-10 | 32 | 58 | µmol/L |
| CREA | women | 11-13 | 34 | 62 | µmol/L |
| CREA | women | 14-17 | 41 | 80 | µmol/L |
| CREA | women | 18-125 | 50 | 90 | µmol/L |
| CREA | men | 5-8 | 26 | 49 | µmol/L |
| CREA | men | 9-10 | 31 | 59 | µmol/L |
| CREA | men | 11-13 | 39 | 68 | µmol/L |
| CREA | men | 14-17 | 52 | 93 | µmol/L |
| CREA | men | 18-125 | 60 | 105 | µmol/L |
| CRP | | | | 10 | mg/L |

| Variable | Sex | Age (years) | Lower boundary | Upper boundary | Unit |
|---|---|---|---|---|---|
| DIMER | | 0-55 | | 50 | FEU/L |
| DIMER | | 56-65 | | 60 | FEU/L |
| DIMER | | 66-75 | | 70 | FEU/L |
| DIMER | | 76-125 | | 80 | FEU/L |
| ECVBE | | | -3 | 3 | |
| EOS | | 0-13 | 0.05 | 0.7 | $10^9$/L |
| EOS | | 14-17 | 0.03 | 0.6 | $10^9$/L |
| ERY | women | 3.94 | 5.16 | | $10^{12}$/L |
| ERY | men | 4.25 | 5.71 | | $10^{12}$/L |
| FERRITIN | | 0-13 | 7 | 140 | µg/L |
| FERRITIN | | 14-125 | 12 | 300 | µg/L |
| GGT | | 0-17 | 10 | 45 | U/L |
| GGT | women | 18-39 | 10 | 45 | U/L |
| GGT | women | 40-125 | 10 | 75 | U/L |
| GGT | men | 18-39 | 10 | 80 | U/L |
| GGT | men | 40-125 | 15 | 115 | U/L |
| GLU | | | 4.2 | 6.3 | mmol/L |
| HAPTO | | 0-13 | 0.05 | 0.5 | g/L |
| HAPTO | | 14-49 | 0.35 | 1.85 | g/L |
| HAPTO | | 50-125 | 0.47 | 2.05 | g/L |
| HB | | 0-1 | 6 8. | 3 | mmol/L |
| HB | | 2-11 | 6.5 | 8.9 | mmol/L |
| HB | | 12-17 | 6.6 | 9.9 | mmol/L |
| HB | women | 18-125 | 7.3 | 9.5 | mmol/L |
| HB | men | 18-125 | 8.3 | 10.5 | mmol/L |
| HBA1C | | | | 48 | mmol/mol |
| HCO3 | | | 22 | 27 | mmol/L |
| HDL | | | 1 | | mmol/L |
| INR | | 0 | 1.3 | | ratio |
| INR | | 1-125 | 1.2 | | ratio |
| JERN | | 0-4 | 5 | 23 | µmol/L |
| JERN | women | 5-10 | 8 | 29 | µmol/L |
| JERN | women | 11-17 | 6 | 33 | µmol/L |
| JERN | men | 5-10 | 6 | 30 | µmol/L |
| JERN | men | 11-17 | 8 | 32 | µmol/L |
| JERN | | 18-125 | 9 | 34 | µmol/L |
| K | | 0-0 | 3.5 | 6.1 | mmol/L |
| K | | 1-4 | 3.3 | 4.6 | mmol/L |
| K | | 5-17 | 3.3 | 4.6 | mmol/L |

| Variable | Sex | Age (years) | Lower boundary | Upper boundary | Unit |
|---|---|---|---|---|---|
| K | | 18-125 | 3.5 | 4.4 | mmol/L |
| KBGLU | | | 4.2 | 6.3 | mmol/L |
| LDH | | 0-3 | 155 | 450 | U/L |
| LDH | | 4-4 | 100 | 345 | U/L |
| LDH | | 5-13 | 157 | 327 | U/L |
| LDH | | 14-17 | 121 | 271 | U/L |
| LDH | | 18-69 | 105 | 205 | U/L |
| LDH | | 70-125 | 115 | 255 | U/L |
| LDL | | | | 3 | mmol/L |
| LEU | | 0-1 | 6.2 | 16.2 | $10^9$/L |
| LEU | | 2-11 | 4.5 | 12.5 | $10^9$/L |
| LEU | | 12-17 | 4.4 | 10.5 | $10^9$/L |
| LEU | | 18-125 | 3.5 | 8.8 | $10^9$/L |
| LYMFO | | 0-5 | 1.83 | 7.85 | $10^9$/L |
| LYMFO | | 6-12 | 1.32 | 4.07 | $10^9$/L |
| LYMFO | | 13-17 | 1.2 | 3.6 | $10^9$/L |
| LYMFO | | 18-125 | 1 | 3.5 | $10^9$/L |
| MCH | | 0-1 | 1.4 | 1.7 | fmol |
| MCH | | 2-5 | 1.5 | 1.7 | fmol |
| MCH | | 6-11 | 1.6 | 1.8 | fmol |
| MCH | | 12-17 | 1.6 | 1.9 | fmol |
| MCH | | 18-125 | 1.7 | 2.1 | fmol |
| MCHC | | 0-17 | 20 | 22 | mmol/L |
| MCHC | | 18-125 | 19.7 | 22.2 | mmol/L |
| MG | | 0-0 | 0.81 | 1.27 | mmol/L |
| MG | | 1-5 | 0.76 | 1 | mmol/L |
| MG | women | 6-13 | 0.73 | 0.93 | mmol/L |
| MG | women | 14-17 | 0.65 | 0.93 | mmol/L |
| MG | men | 6-17 | 0.71 | 0.93 | mmol/L |
| MG | | 18-125 | 0.71 | 0.94 | mmol/L |
| MONO | | 0-17 | 0.21 | 0.77 | $10^9$/L |
| MONO | | 18-125 | 0.2 | 0.8 | $10^9$/L |
| NA | | 0-4 | 137 | 144 | mmol/L |
| NA | | 5-17 | 135 | 147 | mmol/L |
| NA | | 18-125 | 137 | 144 | mmol/L |
| NEUTRO | | 0-13 | 1.6 | 6.7 | $10^9$/L |
| NEUTRO | | 14-17 | 2 | 9.6 | $10^9$/L |
| NEUTRO | | 18-125 | 1.6 | 5.9 | $10^9$/L |
| PHOS | | 0-4 | 1.16 | 1.81 | mmol/L |

| Variable | Sex | Age (years) | Lower boundary | Upper boundary | Unit |
|---|---|---|---|---|---|
| PHOS | women | 5-13 | 1.09 | 1.72 | mmol/L |
| PHOS | women | 14-17 | 0.72 | 1.49 | mmol/L |
| PHOS | women | 18-125 | 0.76 | 1.41 | mmol/L |
| PHOS | men | 5-13 | 1.07 | 1.74 | mmol/L |
| PHOS | men | 14-17 | 0.85 | 1.74 | mmol/L |
| PHOS | men | 18-49 | 0.71 | 1.53 | mmol/L |
| PHOS | men | 50-125 | 0.71 | 1.23 | mmol/L |
| PROCAL | | | | 0.5 | µg/L |
| TRANSJBG | | 0-11 | | 0.41 | |
| TRANSJBG | | 12-17 | | 0.48 | |
| TRANSJBG | women | 18-49 | 0.1 | 0.5 | |
| TRANSJBG | women | 50-125 | 0.15 | 0.5 | |
| TRANSJBG | men | 18-125 | 0.15 | 0.57 | |
| TRIG | | | | 2 | mmol/L |
| TSH | | 0 | 0.73 | 8.92 | $10^{-3}$ IU/L |
| TSH | | 1-5 | 0.69 | 5.89 | $10^{-3}$ IU/L |
| TSH | women | 6-12 | 1.07 | 5.54 | $10^{-3}$ IU/L |
| TSH | women | 13-17 | 0.66 | 5.1 | $10^{-3}$ IU/L |
| TSH | men | 6-17 | 1.06 | 5.8 | $10^{-3}$ IU/L |
| TSH | | 18-125 | 0.4 | 4.8 | $10^{-3}$ IU/L |
| eGFR | | 0-1 | | 70 | mL/min |
| eGFR | | 2-16 | | 80 | mL/min |

## S6

*S6 Most features in the dataset belong to the domain of continuous positive data. This datatype is typically Log-normally distributed.[29] Features that visually resembled log-normal distributed data have therefore been transformed to resemble the Gaussian distribution using the Log-transformation. This table shows the variable name and the transformation applied to it, where Naïve corresponds to no transformation and Log is the natural logarithm.*

| Variable | Data Transformation | Variable | Data Transformation |
|---|---|---|---|
| ABAS | Naïve | INR | Log |
| ABGLU | Log | JERN | Log |
| ABPH | Naïve | K | Log |
| ALAT | Log | KBGLU | Log |
| ALB | Log | LDH | Log |
| AMYL | Log | LDL | Log |
| APTT | Log | LEU | Log |
| ASAT | Log | LYMFO | Log |
| BAS | Naïve | MBAS | Naïve |
| BASO | Log | MCH | Naïve |
| BILI | Log | MCHC | Naïve |
| BILIKON | Log | MG | Log |
| BILIUK | Log | MONO | Log |
| CA | Naïve | NA | Naïve |
| CAI | Naïve | NEUTRO | Log |
| CARB | Log | PHOS | Log |
| CHOL | Log | PROCAL | Log |
| CL | Naïve | TRANSJBG | Naïve |
| CREA | Log | TRIG | Log |
| CRP | Log | TSH | Log |
| DIMER | Log | eGFR | Log |
| ECVBE | Naïve | age | Naïve |
| EOS | Log | BMI | Naïve |
| ERY | Naïve | DiastolicBP | Naïve |
| FERRIITIN | Log | SystolicBP | Naïve |
| GGT | Log | O2Therapy | Naïve |
| GLU | Log | HR | Naïve |
| HAPTO | Log | RespFreq | Naïve |

| Variable | Data Transformation | Variable | Data Transformation |
|---|---|---|---|
| HB | Naïve | Saturation | Naïve |
| HBA1C | Log | Temperature | Naïve |
| HCO3 | Naïve | Weight | Naïve |
| HDL | Log | EWS | Naïve |

**S7-14**

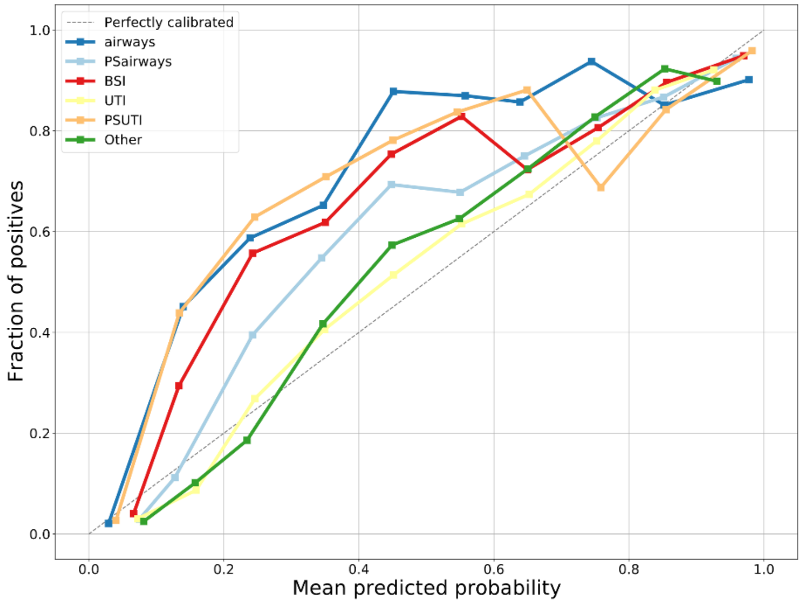


(a) Naive SVM

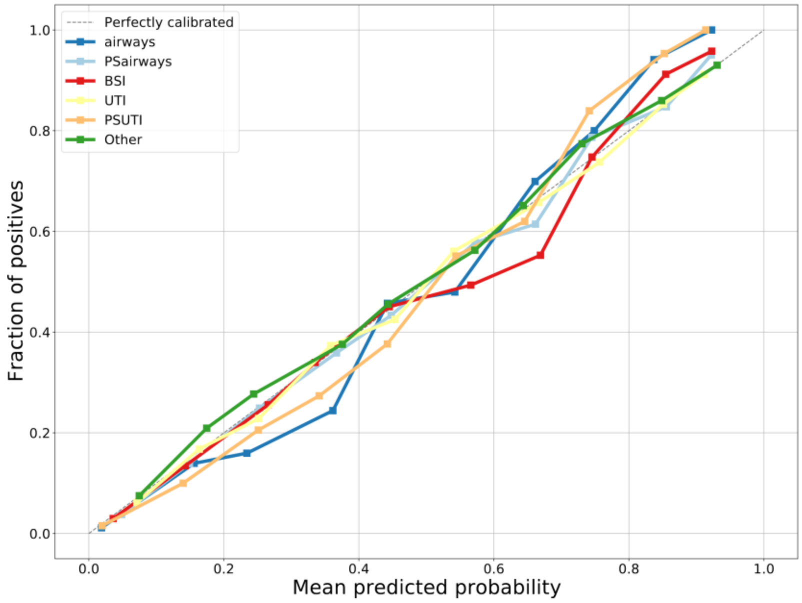


(b) VA calibrated SVM

*S7 Micro averaged calibration curves for the Support Vector Machine (SVM), showing the mean predicted probability vs. the fraction of positives. The blue lines represent airway infections and possible airway infections (PSairways). The red line represents blood stream infections (BSI). The yellow lines represent urinary tract infections (UTI) and possible urinary tract infections (PSUTI). The green line represents other types of infection.*

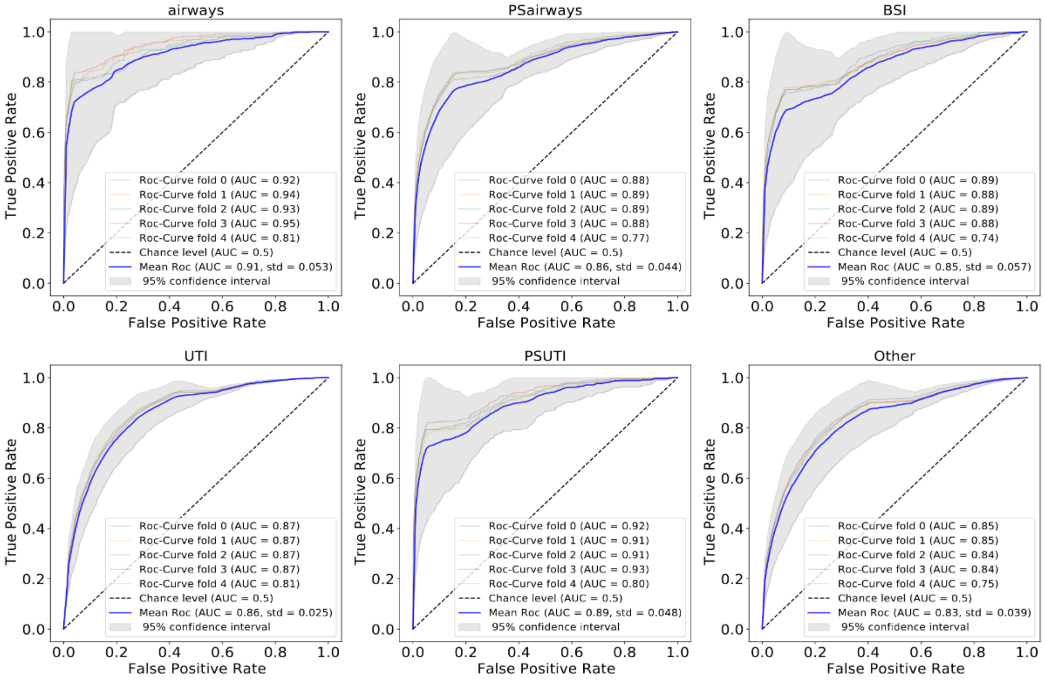


(a)

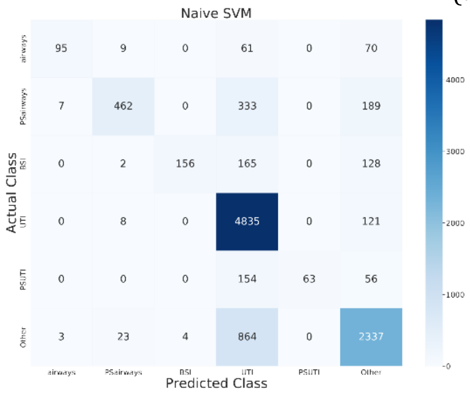


(b)

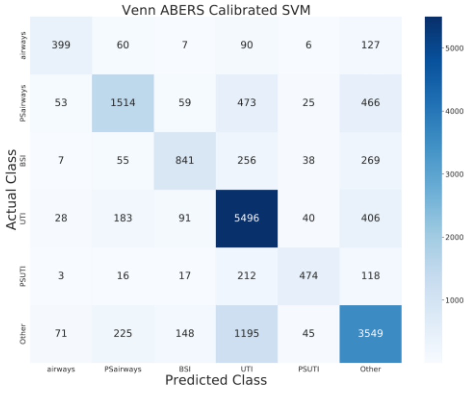


(c)

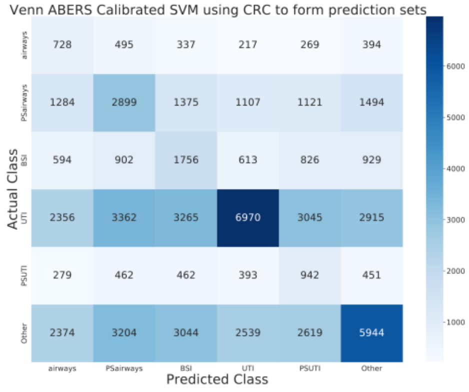


(d)

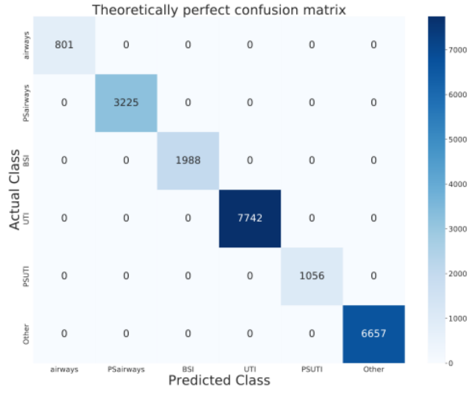


(e)

*S8 Overview of the Support Vector Machine (SVM) performance on the test sets. (a) Receiver operating characteristics (ROC) for each infection class using 5-fold cross validation. (b), (c) (d) Confusion matrices for the SVM in its naïve, VA calibrated, and CRC-VA calibrated from respectively. (e) is the theoretically perfect confusion matrix given the setup. The confusion matrices are summed across the 5-fold cross validation.*

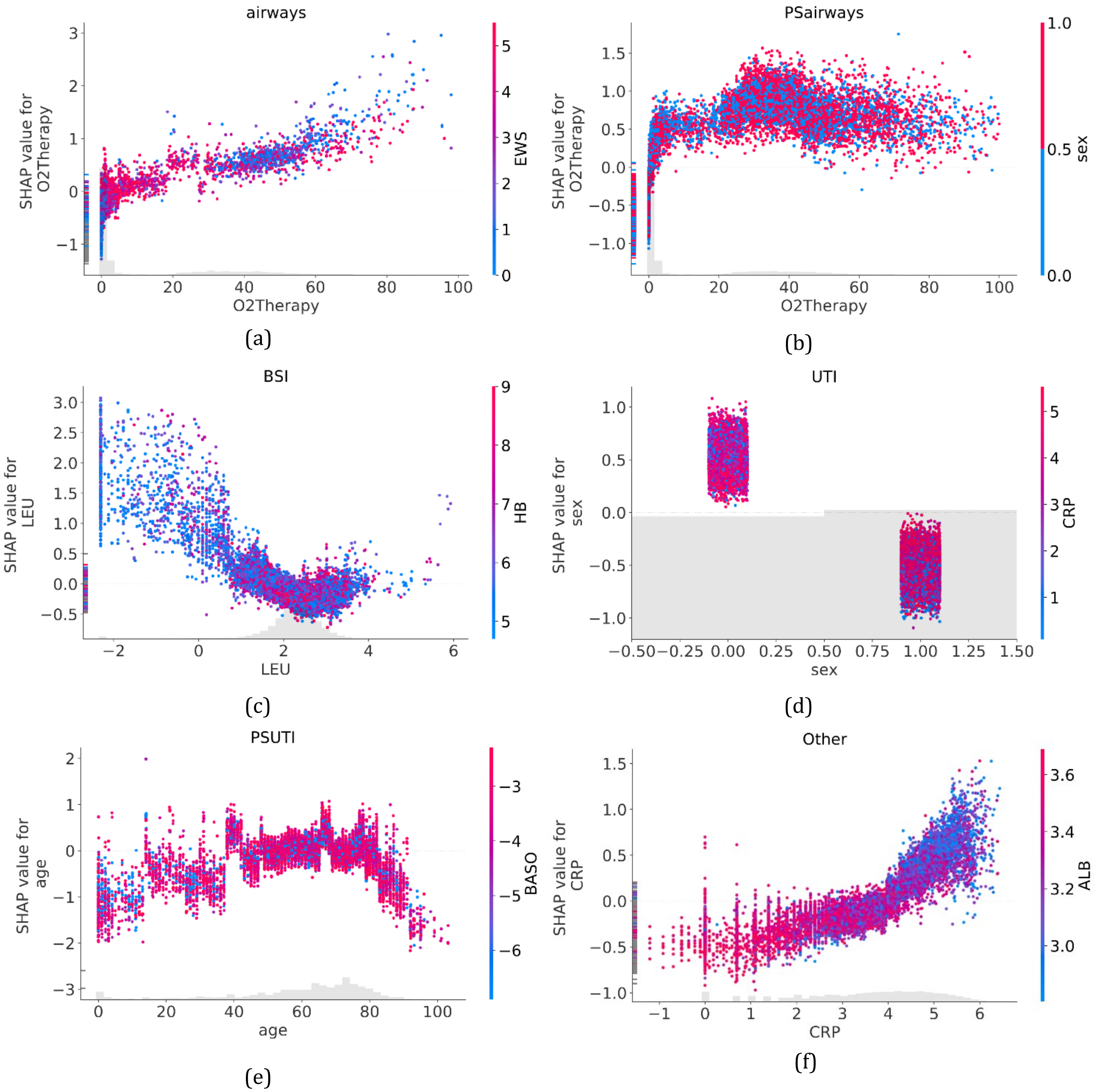


*S9 SHAP summary plots depicting the relationship between the most important feature value (determined by SHAP-feature importance) and the SHAP-value for that feature, as a scatter plot. The points are colored by the second most important feature (determined by SHAP-feature importance). The SHAP-value is equivalent to the log-odds of predicting the class. In (b) sex is encoded categorically as, male=1, female=0. airways: airway infection, BSI: blood stream infection, UTI: urine tract infection, PS: Organisms that are usually not pathogens and common contaminants, O2Therapy: O2 Therapy, EWS: Early warning score, age: patient age in years, LEU: leukocyte count, ALB: albumin, HB: hemoglobin, CRP: C-reactive protein.*

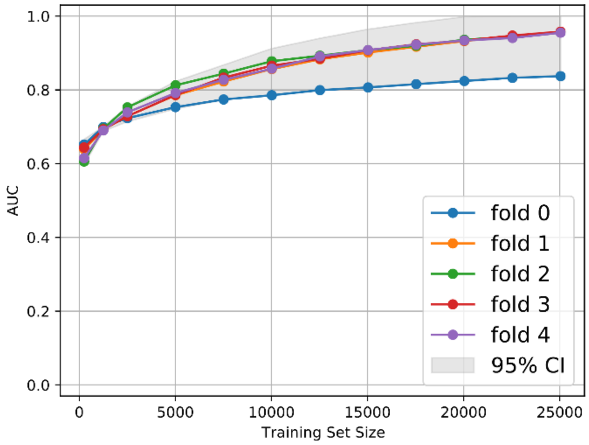


(a) AUC

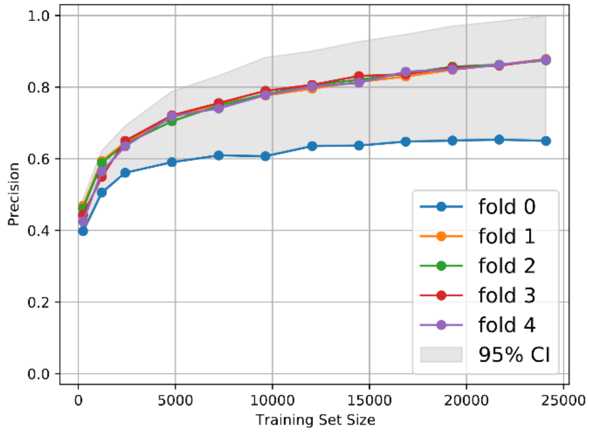


(a) F1-Score

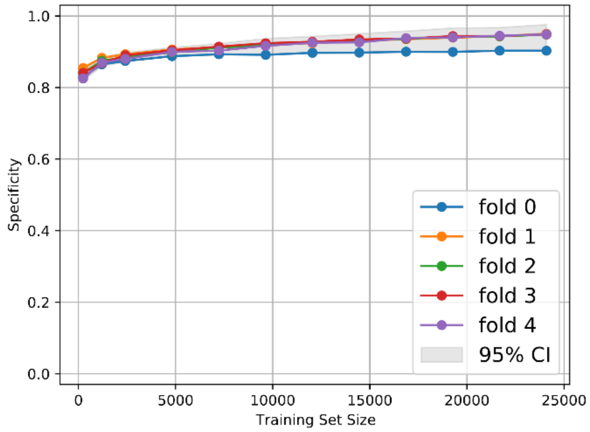


(c) Specificity

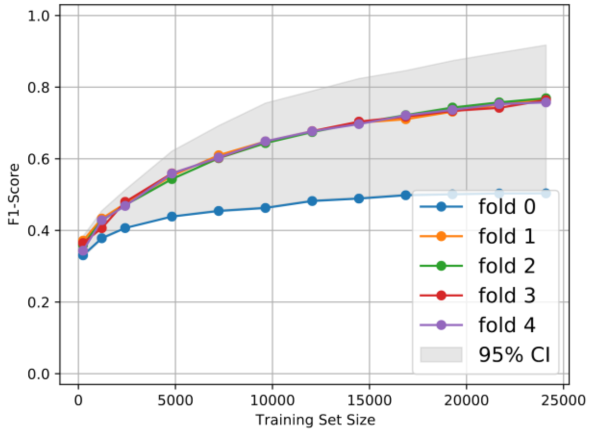


(b) Precision

*S10 Micro averaged performance metrics of the XGBoost model plotted vs. the size of the training dataset, when the model is trained with different training dataset sizes.*

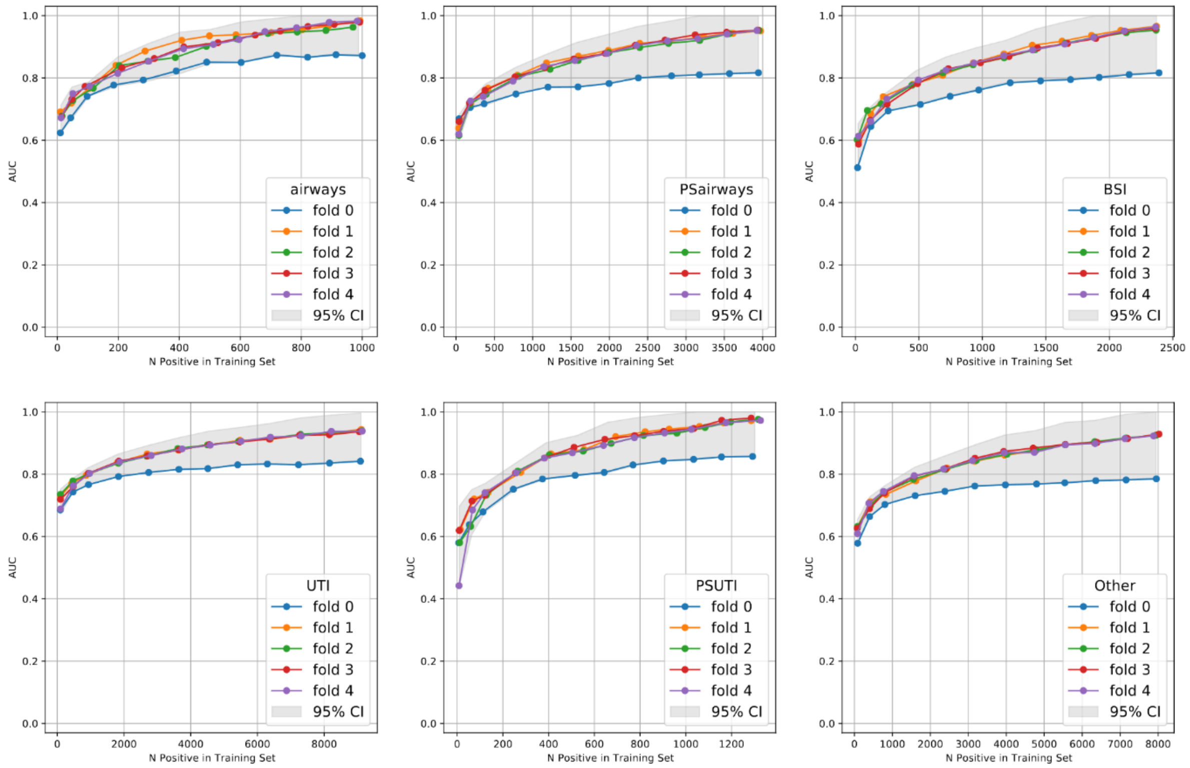


*S11 AUC of the XGBoost model plotted vs. the number of positives in the training dataset for each infection class, when the model is trained with different training dataset sizes. Airways is airway infections, BSI is blood stream infections, UTI is urine tract infections, PS denotes organisms that are usually not pathogens and common contaminants, and the other class is an ensemble of other types of infection.*

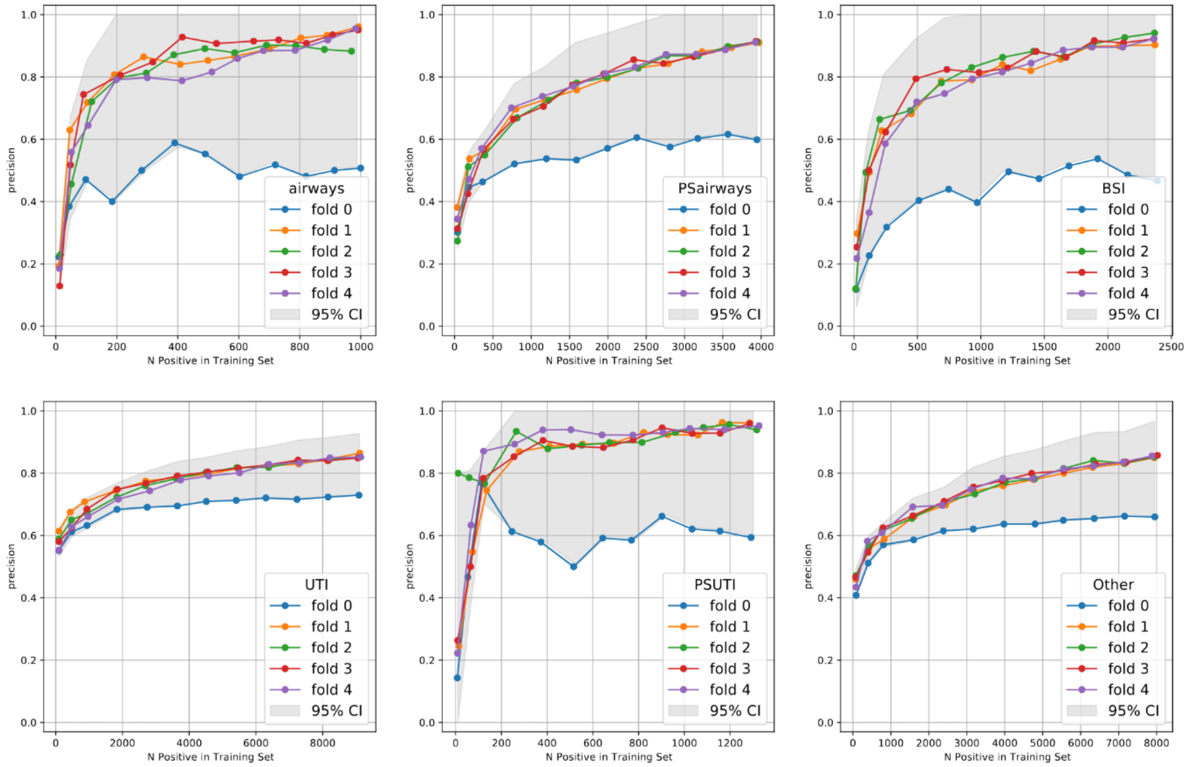


*S12 Precision of the XGBoost model plotted vs. the number of positives in the training dataset for each infection class, when the model is trained with different training dataset sizes. Airways is airway infections, BSI is blood stream infections, UTI is urine tract infections, PS denotes organisms that are usually not pathogens and common contaminants, and the other class is an ensemble of other types of infection.*

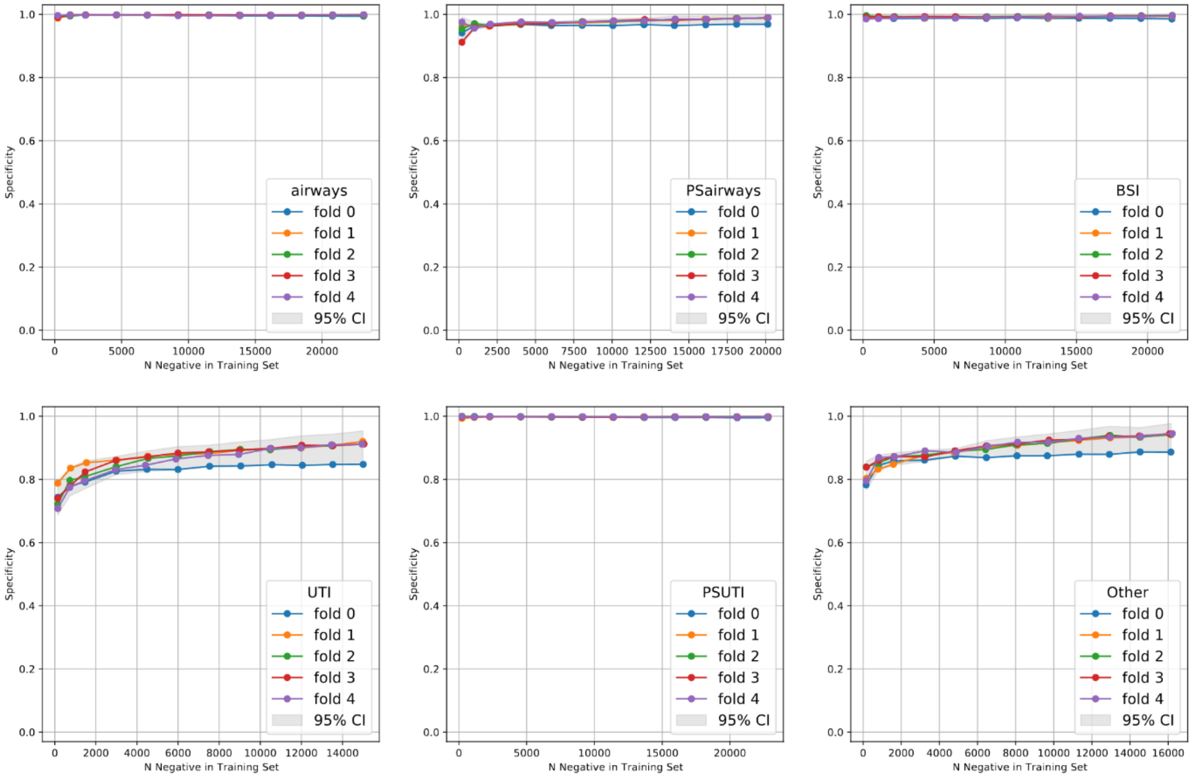


*S13 Specificity of the XGBoost model plotted vs. the number of positives in the training dataset for each infection class, when the model is trained with different training dataset sizes. Airways is airway infections, BSI is blood stream infections, UTI is urine tract infections, PS denotes organisms that are usually not pathogens and common contaminants, and the other class is an ensemble of other types of infection.*

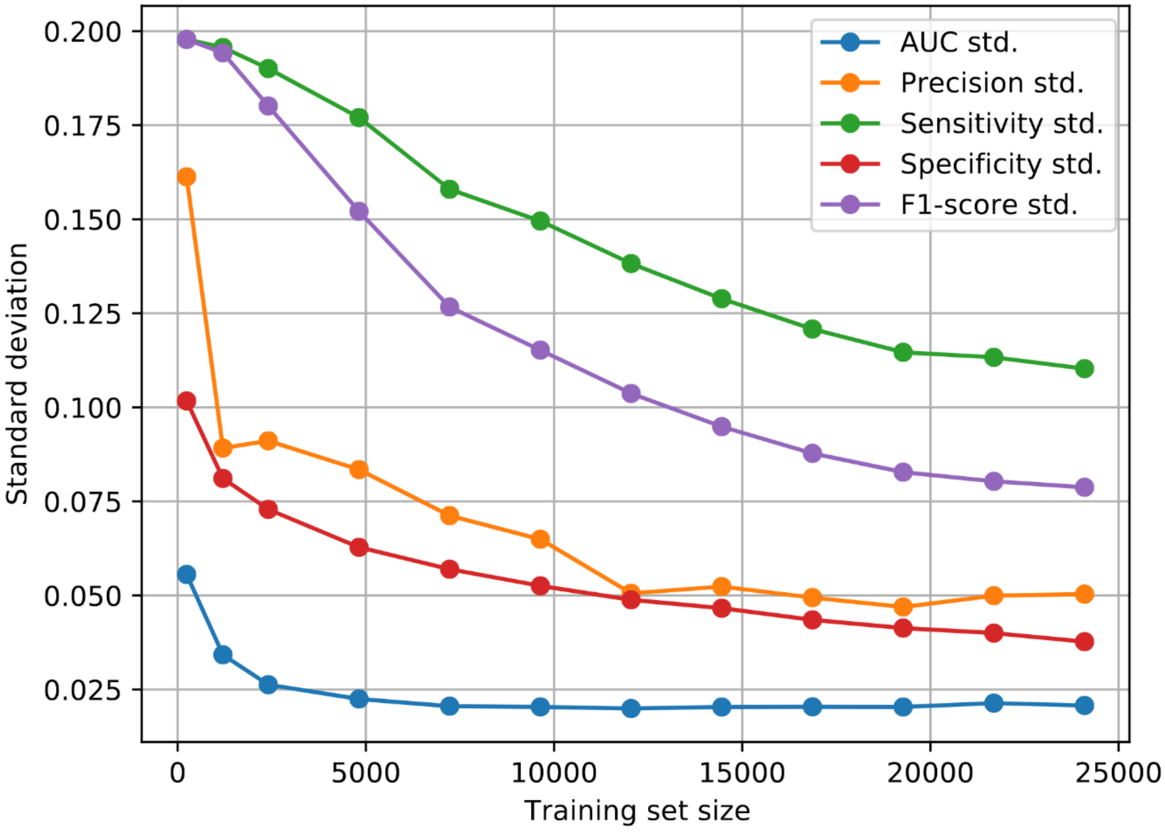


*S 14 Standard deviation of micro averaged performance metrics derived from the XGBoost model vs. training set size.*

## S15

*S15 Table showing the different bacterial species pr. ML label. Airways: airway infections, BSI: Blood stream infections, UTI: Urine tract infections, PS: organisms that are usually not pathogens and common contaminants.*

| Bacteria | Bodily location | ML label | Bacteria | Bodily location | ML label |
|---|---|---|---|---|---|
| Achromobacter spanius | BAL | airways | Acinetobacter junii | BAL | airways |
| Burkholderia cenocepacia | BAL | airways | Citrobacter freundii complex | BAL | airways |
| Citrobacter koseri | BAL | airways | Enterobacter aerogenes | BAL | airways |
| Enterobacter cloacae complex | BAL | airways | Enterococcus faecalis | BAL | airways |
| Enterococcus faecium | BAL | airways | Escherichia coli | BAL | airways |
| Hemolytic streptococci gr. A | BAL | airways | Klebsiella pneumoniae complex | BAL | airways |
| Pandoraea apista | BAL | PSairways | Pseudomonas aeruginosa | BAL | airways |
| Pseudomonas aeruginosa mucoid | BAL | airways | Pseudomonas aeruginosa non mucoid | BAL | airways |
| Serratia marcescens complex | BAL | airways | Stenotrophomonas maltophilia | BAL | PSairways |
| Achromobacter xylosoxidans | Expectorate | PSairways | Acinetobacter baumanii complex | Expectorate | airways |
| Acinetobacter johnsonii | Expectorate | airways | Acinetobacter junii | Expectorate | airways |
| Acinetobacter species | Expectorate | PSairways | Burkholderia species | Expectorate | airways |
| Candida glabrata | Expectorate | PSairways | Citrobacter freundii complex | Expectorate | PSairways |
| Citrobacter koseri | Expectorate | PSairways | Enterobacter aerogenes | Expectorate | PSairways |
| Enterobacter cloacae complex | Expectorate | PSairways | Enterococcus faecalis | Expectorate | PSairways |
| Enterococcus faecium | Expectorate | PSairways | Escherichia coli | Expectorate | PSairways |
| Haemophilus influenzae | Expectorate | airways | Hemolytic streptococci gr. A | Expectorate | airways |
| Klebsiella oxytoca complex | Expectorate | PSairways | Klebsiella pneumoniae complex | Expectorate | PSairways |
| Morganella morganii | Expectorate | airways | Pseudomonas aeruginosa | Expectorate | PSairways |
| Pseudomonas aeruginosa mucoid | Expectorate | PSairways | Pseudomonas aeruginosa non mucoid | Expectorate | PSairways |
| Pseudomonas fluorescens gruppen | Expectorate | PSairways | Pseudomonas putida group | Expectorate | PSairways |
| Pseudomonas species | Expectorate | PSairways | Serratia liquefaciens complex | Expectorate | PSairways |
| Serratia marcescens complex | Expectorate | PSairways | Staphylococcus aureus | Expectorate | PSairways |
| Stenotrophomonas maltophilia | Expectorate | PSairways | Acinetobacter lwoffii | Pleural effusion | airways |
| Citrobacter freundii complex | Pleural effusion | airways | Enterobacter cloacae complex | Pleural effusion | airways |
| Enterococcus faecalis | Pleural effusion | airways | Enterococcus faecium | Pleural effusion | PSairways |
| Enterococcus gallinarum | Pleural effusion | PSairways | Escherichia coli | Pleural effusion | airways |
| Hemolytic streptococci gr. A | Pleural effusion | airways | Klebsiella oxytoca complex | Pleural effusion | airways |
| Klebsiella pneumoniae complex | Pleural effusion | airways | Pseudomonas aeruginosa | Pleural effusion | airways |
| Pseudomonas stutzeri gruppen | Pleural effusion | PSairways | Staphylococcus aureus | Pleural effusion | airways |

| **Bacteria** | **Bodily location** | **ML label** | **Bacteria** | **Bodily location** | **ML label** |
|---|---|---|---|---|---|
| Stenotrophomonas maltophilia | Pleural effusion | PSairways | Acinetobacter baumanii complex | Tracheal secretions | airways |
| Acinetobacter johnsonii | Tracheal secretions | PSairways | Acinetobacter junii | Tracheal secretions | PSairways |
| Acinetobacter species | Tracheal secretions | PSairways | Citrobacter freundii complex | Tracheal secretions | PSairways |
| Citrobacter koseri | Tracheal secretions | PSairways | Enterobacter aerogenes | Tracheal secretions | PSairways |
| Enterobacter cloacae complex | Tracheal secretions | PSairways | Enterococcus faecalis | Tracheal secretions | PSairways |
| Enterococcus faecium | Tracheal secretions | PSairways | Escherichia coli | Tracheal secretions | PSairways |
| Hemolytic streptococci gr. A | Tracheal secretions | airways | Klebsiella oxytoca complex | Tracheal secretions | PSairways |
| Klebsiella pneumoniae complex | Tracheal secretions | PSairways | Pseudomonas aeruginosa | Tracheal secretions | airways |
| Pseudomonas aeruginosa mucoid | Tracheal secretions | airways | Pseudomonas aeruginosa non mucoid | Tracheal secretions | airways |
| Serratia liquefaciens complex | Tracheal secretions | PSairways | Serratia marcescens complex | Tracheal secretions | PSairways |
| Staphylococcus aureus | Tracheal secretions | PSairways | Stenotrophomonas maltophilia | Tracheal secretions | PSairways |
| Stenotrophomonas species | Tracheal secretions | PSairways | Enterococcus faecium | Blood (blood culture flask) | BSI |
| Escherichia coli | Blood (blood culture flask) | BSI | Klebsiella pneumoniae complex | Blood (blood culture flask) | BSI |
| Citrobacter koseri | Blood from artery (flask) | BSI | Enterobacter aerogenes | Blood from artery (flask) | BSI |
| Enterobacter cloacae complex | Blood from artery (flask) | BSI | Enterococcus faecalis | Blood from artery (flask) | BSI |
| Enterococcus faecium | Blood from artery (flask) | BSI | Escherichia coli | Blood from artery (flask) | BSI |
| Klebsiella oxytoca complex | Blood from artery (flask) | BSI | Klebsiella pneumoniae complex | Blood from artery (flask) | BSI |
| Pseudomonas aeruginosa | Blood from artery (flask) | BSI | Acinetobacter baumanii complex | Blood from catheter (flask) | BSI |
| Acinetobacter lwoffii | Blood from catheter (flask) | BSI | Acinetobacter species | Blood from catheter (flask) | BSI |
| Citrobacter freundii complex | Blood from catheter (flask) | BSI | Citrobacter koseri | Blood from catheter (flask) | BSI |
| Enterobacter aerogenes | Blood from catheter (flask) | BSI | Enterobacter cloacae complex | Blood from catheter (flask) | BSI |

| Bacteria | Bodily location | ML label | Bacteria | Bodily location | ML label |
|---|---|---|---|---|---|
| Enterococcus faecalis | Blood from catheter (flask) | BSI | Enterococcus faecium | Blood from catheter (flask) | BSI |
| Enterococcus gallinarum | Blood from catheter (flask) | BSI | Escherichia coli | Blood from catheter (flask) | BSI |
| Klebsiella oxytoca complex | Blood from catheter (flask) | BSI | Klebsiella pneumoniae complex | Blood from catheter (flask) | BSI |
| Kluyvera cryocrescens | Blood from catheter (flask) | PSBSI | Pseudomonas aeruginosa | Blood from catheter (flask) | BSI |
| Pseudomonas putida group | Blood from catheter (flask) | BSI | Serratia marcescens complex | Blood from catheter (flask) | BSI |
| Staphylococcus aureus | Blood from catheter (flask) | BSI | Stenotrophomonas maltophilia | Blood from catheter (flask) | BSI |
| Acinetobacter baumanii complex | Blood from peripheral vein (flask) | BSI | Acinetobacter species | Blood from peripheral vein (flask) | PSBSI |
| Citrobacter freundii complex | Blood from peripheral vein (flask) | BSI | Citrobacter koseri | Blood from peripheral vein (flask) | BSI |
| Enterobacter aerogenes | Blood from peripheral vein (flask) | BSI | Enterobacter cloacae complex | Blood from peripheral vein (flask) | BSI |
| Enterococcus faecalis | Blood from peripheral vein (flask) | BSI | Enterococcus faecium | Blood from peripheral vein (flask) | BSI |
| Escherichia coli | Blood from peripheral vein (flask) | BSI | Klebsiella oxytoca complex | Blood from peripheral vein (flask) | BSI |
| Klebsiella pneumoniae complex | Blood from peripheral vein (flask) | BSI | Mycobacterium septicum | Blood from peripheral vein (flask) | BSI |
| Proteus vulgaris complex | Blood from peripheral vein (flask) | BSI | Bacteria | Bodily location | ML label |
| Pseudomonas aeruginosa | Blood from peripheral vein (flask) | BSI | Serratia marcescens complex | Blood from peripheral vein (flask) | BSI |
| Staphylococcus aureus | Blood from peripheral vein (flask) | BSI | Staphylococcus species | Blood from peripheral vein (flask) | PSBSI |
| Stenotrophomonas maltophilia | Blood from peripheral vein (flask) | BSI | Acinetobacter baumanii complex | Urine | UTI |

| Bacteria | Bodily location | ML label | Bacteria | Bodily location | ML label |
|---|---|---|---|---|---|
| Citrobacter freundii complex | Urine | UTI | Citrobacter koseri | Urine | UTI |
| Enterobacter cloacae complex | Urine | UTI | Enterococcus faecalis | Urine | UTI |
| Enterococcus faecium | Urine | PSUTI | Escherichia coli | Urine | UTI |
| Klebsiella oxytoca complex | Urine | UTI | Klebsiella pneumoniae complex | Urine | UTI |
| Morganella morganii | Urine | UTI | Pseudomonas aeruginosa | Urine | UTI |
| Stenotrophomonas maltophilia | Urine | PSUTI | Escherichia coli | Urine (catheter) | PSUTI |
| Acinetobacter baumanii complex | Urine (midstream) | UTI | Acinetobacter johnsonii | Urine (midstream) | PSUTI |
| Acinetobacter junii | Urine (midstream) | PSUTI | Acinetobacter ursingii | Urine (midstream) | PSUTI |
| Citrobacter amalonaticus complex | Urine (midstream) | UTI | Citrobacter freundii complex | Urine (midstream) | UTI |
| Citrobacter koseri | Urine (midstream) | UTI | Enterobacter aerogenes | Urine (midstream) | UTI |
| Enterobacter cloacae complex | Urine (midstream) | UTI | Enterococcus avium | Urine (midstream) | PSUTI |
| Enterococcus faecalis | Urine (midstream) | UTI | Enterococcus faecium | Urine (midstream) | PSUTI |
| Enterococcus gallinarum | Urine (midstream) | PSUTI | Enterococcus mundtii | Urine (midstream) | PSUTI |
| Enterococcus raffinosus | Urine (midstream) | PSUTI | Escherichia coli | Urine (midstream) | UTI |
| Gram negative rods | Urine (midstream) | UTI | Hemolytic streptococci gr. A | Urine (midstream) | UTI |
| Klebsiella oxytoca complex | Urine (midstream) | UTI | Klebsiella pneumoniae complex | Urine (midstream) | UTI |
| Klebsiella species | Urine (midstream) | UTI | Morganella morganii | Urine (midstream) | UTI |
| Proteus mirabilis | Urine (midstream) | UTI | Pseudomonas aeruginosa | Urine (midstream) | UTI |
| Pseudomonas aeruginosa mucoid | Urine (midstream) | UTI | Serratia marcescens complex | Urine (midstream) | UTI |
| Staphylococcus aureus | Urine (midstream) | PSUTI | Stenotrophomonas maltophilia | Urine (midstream) | PSUTI |
| Acinetobacter baumanii complex | Urine (nephrostomy catheter) | UTI | Acinetobacter species | Urine (nephrostomy catheter) | PSUTI |
| Acinetobacter ursingii | Urine (nephrostomy catheter) | PSUTI | Citrobacter freundii complex | Urine (nephrostomy catheter) | UTI |

| Bacteria | Bodily location | ML label | Bacteria | Bodily location | ML label |
|---|---|---|---|---|---|
| Citrobacter koseri | Urine (nephrostomy catheter) | UTI | Enterobacter aerogenes | Urine (nephrostomy catheter) | UTI |
| Enterobacter cloacae complex | Urine (nephrostomy catheter) | UTI | Enterococcus avium | Urine (nephrostomy catheter) | PSUTI |
| Enterococcus casseliflavus | Urine (nephrostomy catheter) | PSUTI | Enterococcus faecalis | Urine (nephrostomy catheter) | UTI |
| Enterococcus faecium | Urine (nephrostomy catheter) | PSUTI | Enterococcus gallinarum | Urine (nephrostomy catheter) | PSUTI |
| Enterococcus hirae | Urine (nephrostomy catheter) | PSUTI | Escherichia coli | Urine (nephrostomy catheter) | UTI |
| Klebsiella oxytoca complex | Urine (nephrostomy catheter) | UTI | Klebsiella pneumoniae complex | Urine (nephrostomy catheter) | UTI |
| Non-hemolytic streptococ mitis group | Urine (nephrostomy catheter) | PSUTI | Pseudomonas aeruginosa | Urine (nephrostomy catheter) | UTI |
| Pseudomonas putida group | Urine (nephrostomy catheter) | PSUTI | Serratia marcescens complex | Urine (nephrostomy catheter) | UTI |
| Staphylococcus aureus | Urine (nephrostomy catheter) | PSUTI | Stenotrophomonas maltophilia | Urine (nephrostomy catheter) | PSUTI |
| Acinetobacter baumanii complex | Urine (suprapubic puncture) | UTI | Acinetobacter johnsonii | Urine (suprapubic puncture) | UTI |
| Acinetobacter junii | Urine (suprapubic puncture) | UTI | Citrobacter freundii complex | Urine (suprapubic puncture) | UTI |
| Citrobacter koseri | Urine (suprapubic puncture) | UTI | Enterobacter aerogenes | Urine (suprapubic puncture) | UTI |
| Enterobacter cloacae complex | Urine (suprapubic puncture) | UTI | Enterococcus faecalis | Urine (suprapubic puncture) | UTI |
| Enterococcus faecium | Urine (suprapubic puncture) | UTI | Escherichia coli | Urine (suprapubic puncture) | UTI |
| Klebsiella oxytoca complex | Urine (suprapubic puncture) | UTI | Klebsiella pneumoniae complex | Urine (suprapubic puncture) | UTI |
| Pseudomonas aeruginosa | Urine (suprapubic puncture) | UTI | Serratia marcescens complex | Urine (suprapubic puncture) | UTI |

| Bacteria | Bodily location | ML label | Bacteria | Bodily location | ML label |
|---|---|---|---|---|---|
| Stenotrophomonas maltophilia | Urine (suprapubic puncture) | UTI | Escherichia coli | Urine (suprapubic/ topcatheter) | UTI |
| Citrobacter freundii complex | Urine from J-J catheter | UTI | Enterobacter aerogenes | Urine from J-J catheter | UTI |
| Enterococcus faecalis | Urine from J-J catheter | UTI | Enterococcus faecium | Urine from J-J catheter | PSUTI |
| Escherichia coli | Urine from J-J catheter | UTI | Klebsiella oxytoca complex | Urine from J-J catheter | UTI |
| Klebsiella pneumoniae complex | Urine from J-J catheter | UTI | Pseudomonas aeruginosa | Urine from J-J catheter | UTI |
| Citrobacter freundii complex | Urine from KAD | UTI | Citrobacter koseri | Urine from KAD | UTI |
| Enterobacter aerogenes | Urine from KAD | UTI | Enterobacter cloacae complex | Urine from KAD | UTI |
| Enterococcus faecalis | Urine from KAD | UTI | Enterococcus faecium | Urine from KAD | PSUTI |
| Escherichia coli | Urine from KAD | UTI | Hemolytic streptococci gr. A | Urine from KAD | UTI |
| Klebsiella oxytoca complex | Urine from KAD | UTI | Klebsiella pneumoniae complex | Urine from KAD | UTI |
| Klebsiella species | Urine from KAD | UTI | Proteus mirabilis | Urine from KAD | UTI |
| Pseudomonas aeruginosa | Urine from KAD | UTI | Pseudomonas aeruginosa mucoid | Urine from KAD | UTI |
| Serratia marcescens complex | Urine from KAD | UTI | Stenotrophomonas maltophilia | Urine from KAD | PSUTI |
| Citrobacter freundii complex | Urine from reservoir (Bricker) | UTI | Citrobacter koseri | Urine from reservoir (Bricker) | UTI |
| Enterobacter cloacae complex | Urine from reservoir (Bricker) | UTI | Enterococcus casseliflavus | Urine from reservoir (Bricker) | PSUTI |
| Enterococcus durans | Urine from reservoir (Bricker) | PSUTI | Enterococcus faecalis | Urine from reservoir (Bricker) | UTI |
| Enterococcus faecium | Urine from reservoir (Bricker) | PSUTI | Enterococcus gallinarum | Urine from reservoir (Bricker) | PSUTI |
| Enterococcus raffinosus | Urine from reservoir (Bricker) | PSUTI | Enterococcus species | Urine from reservoir (Bricker) | PSUTI |
| Escherichia coli | Urine from reservoir (Bricker) | UTI | Klebsiella oxytoca complex | Urine from reservoir (Bricker) | UTI |
| Klebsiella pneumoniae complex | Urine from reservoir (Bricker) | UTI | Pseudomonas aeruginosa | Urine from reservoir (Bricker) | UTI |

| Bacteria | Bodily location | ML label | Bacteria | Bodily location | ML label |
|---|---|---|---|---|---|
| Serratia marcescens complex | Urine from reservoir (Bricker) | UTI | Staphylococcus aureus | Urine from reservoir (Bricker) | PSUTI |
| Stenotrophomonas maltophilia | Urine from reservoir (Bricker) | PSUTI | | | |

## S16

*S16 Table of summary statistics (Median, [IQR] and number of observations) grouped by the class of infection (for binary variables only the number of observations is given). Variable describes the name of the feature in the dataset and description is a short description of the feature. Airway is airway infections; BSI is blood stream infections; and UTI is urinary tract infections; PS denotes organisms that are usually not pathogens and common contaminants. BAS is the percentage of biochemical test results outside the normal range; ABAS is the percentage of available biochemical test results outside the normal range; MBAS is the percentage of selected biochemical test results outside the normal range (see S2 and S5)*

| Variable | Description | Unit | Airways | PSAirways | BSI | UTI | PSUTI | Other |
|---|---|---|---|---|---|---|---|---|
| ABAS | Adjusted biochemical abnormality score | % | 57.14 [46.15, 66.67] 1228 | 57.14 [45.45, 66.67] 4909 | 57.14 [45.45, 68.42] 2967 | 50 [35.29, 61.54] 11430 | 57.14 [47.06, 68.75] 1622 | 58.33 [46.15, 69.23] 9920 |
| ABGLU | Glucose arterie blood | mmol/L | 7.4 [6.29, 8.80] 14 | 9.3 [7.33, 10.94] 39 | 8.95 [6.88, 11.12] 18 | 7.5 [6.20, 8.90] 84 | 7.92 [6.75, 8.53] 8 | 8.46 [7.00, 13.10] 21 |
| ABPH | Arterial blood pH | pH | 7.41 [7.35, 7.44] 50 | 7.41 [7.34, 7.46] 184 | 7.42 [7.35, 7.45] 67 | 7.42 [7.37, 7.46] 242 | 7.44 [7.41, 7.47] 45 | 7.41 [7.35, 7.46] 244 |
| ALAT | Alanine aminotransferase | U/L | 31 [20.00, 47.50] 511 | 28 [18.00, 46.00] 2035 | 29 [18.00, 48.00] 1217 | 23 [16.00, 36.00] 4215 | 25 [16.00, 39.00] 587 | 29 [18.00, 47.00] 3689 |
| ALB | Albumin | g/L | 26 [21.00, 32.00] 700 | 26 [22.00, 31.00] 2624 | 26.5 [22.00, 31.00] 1405 | 31 [26.00, 36.00] 6517 | 25.75 [21.00, 30.00] 848 | 25 [21.00, 30.00] 5038 |
| AMYL | Amylase | U/L | 61 [40.00, 88.00] 382 | 61 [41.00, 92.00] 1166 | 56 [37.00, 79.00] 502 | 60 [42.00, 83.00] 1513 | 50 [29.00, 81.50] 167 | 56 [35.00, 88.00] 1792 |
| APTT | Activate partial thromboplastin time | s | 37 [31.00, 45.00] 264 | 32 [28.00, 40.00] 576 | 33 [27.00, 39.00] 260 | 30 [27.00, 34.00] 1268 | 32.5 [28.00, 41.00] 97 | 33 [29.00, 40.00] 883 |
| ASAT | Aspartate transaminase | U/L | 49 [33.50, 111.50] 227 | 56.5 [30.00, 114.75] 502 | 54 [32.00, 114.00] 201 | 29 [21.00, 49.00] 626 | 41 [27.50, 67.75] 78 | 56 [29.00, 123.00] 769 |
| BAS | Biochemical abnormality score | % | 16 [12.00, 20.00] 1228 | 16 [10.00, 20.00] 4910 | 14 [10.00, 18.00] 2968 | 12 [8.00, 18.00] 11430 | 14 [10.00, 18.00] 1622 | 14 [10.00, 18.00] 9920 |
| BASO | Basophilocyte count | $10^9$/L | 0.03 [0.01, 0.06] 510 | 0.03 [0.02, 0.06] 2400 | 0.03 [0.01, 0.05] 911 | 0.03 [0.01, 0.06] 5711 | 0.04 [0.02, 0.06] 596 | 0.03 [0.01, 0.06] 3235 |

| Variable | Description | Unit | Airways | PSAirways | BSI | UTI | PSUTI | Other |
|---|---|---|---|---|---|---|---|---|
| BILI | Bilirubin | µmol/L | 10<br>[6.00, 20.00]<br>563 | 9<br>[6.00, 16.00]<br>2164 | 12<br>[7.00, 23.61]<br>1499 | 8<br>[5.00, 14.00]<br>3915 | 8.5<br>[5.00, 15.00]<br>642 | 11<br>[6.00, 24.00]<br>4313 |
| BILIKON | Conjugated bilirubin | µmol/L | 101<br>[8.10, 194.75]<br>4 | 9<br>[2.00, 84.00]<br>33 | 13.5<br>[9.00, 18.75]<br>30 | 2<br>[2.00, 5.25]<br>104 | 5<br>[3.00, 90.00]<br>5 | 11<br>[6.00, 86.00]<br>77 |
| BILIUK | Unconjugated bilirubin | µmol/L | 101.5<br>[59.75, 144.25]<br>4 | 61<br>[9.00, 126.00]<br>25 | 23<br>[6.00, 98.00]<br>29 | 8<br>[5.00, 40.00]<br>61 | 4<br>[3.00, 24.00]<br>5 | 36<br>[11.00, 106.50]<br>68 |
| BMI | Body mass index | $kg/m^2$ | 18.31<br>[13.50, 27.02]<br>10 | 25.52<br>[20.36, 28.70]<br>32 | 20.21<br>[16.70, 24.81]<br>57 | 25.26<br>[21.36, 28.66]<br>242 | 23.95<br>[18.57, 26.83]<br>24 | 23.75<br>[17.82, 27.49]<br>126 |
| CA | Calcium | mmol/L | 2.23<br>[2.10, 2.34]<br>211 | 2.19<br>[2.07, 2.32]<br>823 | 2.19<br>[2.06, 2.34]<br>320 | 2.28<br>[2.16, 2.39]<br>1468 | 2.24<br>[2.13, 2.35]<br>158 | 2.21<br>[2.09, 2.34]<br>770 |
| CAI | Ionized calcium | mmol/L | 1.2<br>[1.13, 1.26]<br>197 | 1.21<br>[1.14, 1.27]<br>772 | 1.22<br>[1.16, 1.27]<br>757 | 1.23<br>[1.18, 1.28]<br>3003 | 1.22<br>[1.16, 1.29]<br>435 | 1.2<br>[1.14, 1.26]<br>1687 |
| CARB | Carbamide | mmol/L | 8.5<br>[5.50, 14.00]<br>947 | 8.25<br>[5.50, 13.50]<br>3290 | 8.95<br>[5.70, 15.30]<br>1499 | 6.9<br>[4.50, 11.10]<br>5925 | 11.5<br>[6.30, 20.00]<br>801 | 8.4<br>[5.20, 14.60]<br>5446 |
| CHOL | Cholesterol | mmol/L | 4.25<br>[3.63, 5.55]<br>28 | 3.9<br>[3.33, 5.25]<br>147 | 3.85<br>[3.10, 4.70]<br>104 | 4.4<br>[3.60, 5.30]<br>1010 | 4<br>[3.50, 5.30]<br>27 | 3.7<br>[2.90, 4.60]<br>247 |
| CL | Chloride | mmol/L | 105.42<br>[101.00, 110.38]<br>76 | 105<br>[101.50, 109.00]<br>183 | 105<br>[100.00, 108.64]<br>115 | 105<br>[102.00, 107.00]<br>545 | 104.29<br>[102.00, 106.00]<br>46 | 105.5<br>[102.00, 109.00]<br>301 |
| CREA | Creatinine | µmol/L | 81<br>[55.50, 128.50]<br>1157 | 80<br>[56.00, 129.00]<br>4605 | 82<br>[58.00, 136.00]<br>2689 | 72<br>[54.00, 107.00]<br>10415 | 94<br>[61.00, 180.75]<br>1538 | 79<br>[56.00, 129.00]<br>8967 |
| CRP | C-reactive protein | mg/L | 57<br>[17.00, 139.00]<br>1165 | 56<br>[18.00, 131.00]<br>4557 | 45<br>[11.00, 116.88]<br>2602 | 21<br>[6.00, 58.00]<br>9505 | 49.5<br>[16.00, 109.00]<br>1495 | 67<br>[22.00, 143.00]<br>8865 |
| DIMER | Fibrin D-Dimer | FEU/L | 5.1<br>[1.85, 11.95]<br>201 | 4.1<br>[1.38, 9.20]<br>343 | 3.2<br>[1.30, 6.65]<br>115 | 1.3<br>[0.60, 4.25]<br>319 | 2.5<br>[1.10, 6.30]<br>51 | 3.5<br>[1.40, 7.30]<br>463 |
| DiastolicBP | Diastolic blood pressure | mmHg | 60.41<br>[54.57, 68.82]<br>1149 | 63.44<br>[55.82, 72.19]<br>4678 | 66.5<br>[57.39, 75.00]<br>2698 | 68.83<br>[60.58, 77.75]<br>9999 | 67.8<br>[60.18, 76.73]<br>1539 | 65.49<br>[56.75, 74.50]<br>9197 |
| ECVBE | Base excess Ecv | | 1.1<br>[-2.52, 4.22]<br>99 | 0.98<br>[-2.60, 4.11]<br>252 | -1.2<br>[-4.95, 1.70]<br>125 | -0.37<br>[-2.67, 2.00]<br>546 | 1.64<br>[-0.90, 3.70]<br>73 | -0.25<br>[-4.00, 3.00]<br>419 |
| EOS | Eosinophil count | $10^9/L$ | 0.08<br>[0.01, 0.24]<br>510 | 0.1<br>[0.02, 0.26]<br>2400 | 0.04<br>[0.01, 0.17]<br>911 | 0.1<br>[0.03, 0.22]<br>5713 | 0.12<br>[0.03, 0.25]<br>596 | 0.1<br>[0.02, 0.25]<br>3236 |
| ERY | Erythrocyte count | $10^{12}/L$ | 3.59<br>[3.12, 4.23]<br>108 | 3.7<br>[3.12, 4.31]<br>554 | 3.44<br>[3.04, 3.89]<br>187 | 3.96<br>[3.41, 4.47]<br>1663 | 3.37<br>[2.97, 3.90]<br>153 | 3.5<br>[3.00, 4.15]<br>703 |
| EWS | Early warning score | Integer | 2.67<br>[1.00, 4.36]<br>342 | 2<br>[1.00, 4.00]<br>2184 | 1.33<br>[0.50, 3.00]<br>1789 | 1<br>[0.00, 2.00]<br>7231 | 1.33<br>[0.50, 2.50]<br>1233 | 1.2<br>[0.50, 2.67]<br>6144 |

| Variable | Description | Unit | Airways | PSAirways | BSI | UTI | PSUTI | Other |
|---|---|---|---|---|---|---|---|---|
| FERRITIN | Ferritin | µg/L | 452<br>[243.00, 664.00]<br>113 | 482<br>[213.00, 1100.00]<br>233 | 322<br>[130.00, 754.50]<br>183 | 282<br>[120.00, 622.00]<br>717 | 469<br>[282.00, 903.00]<br>57 | 366<br>[155.00, 767.00]<br>541 |
| GGT | Gamma glutamyl transferase | U/L | 150<br>[69.25, 320.25]<br>46 | 62<br>[31.00, 142.00]<br>133 | 114.5<br>[48.00, 186.88]<br>88 | 47<br>[22.00, 148.25]<br>444 | 136<br>[23.00, 280.00]<br>21 | 94<br>[37.00, 213.50]<br>275 |
| GLU | Glucose | mmol/L | 7.4<br>[5.60, 9.60]<br>199 | 6.9<br>[5.60, 9.00]<br>1028 | 6.6<br>[5.50, 8.60]<br>991 | 6.5<br>[5.55, 8.20]<br>3561 | 6.5<br>[5.60, 8.40]<br>646 | 6.7<br>[5.60, 8.60]<br>2132 |
| HAPTO | Haptoglobin | g/L | 0.96<br>[0.55, 1.61]<br>26 | 0.58<br>[0.10, 2.20]<br>81 | 1.14<br>[0.26, 2.21]<br>62 | 1.19<br>[0.10, 2.21]<br>193 | 1.21<br>[0.11, 2.16]<br>36 | 1.06<br>[0.10, 2.36]<br>194 |
| HB | Hemoglobin | mmol/L | 6.2<br>[5.40, 7.20]<br>1201 | 6.4<br>[5.50, 7.50]<br>4734 | 6.1<br>[5.40, 7.05]<br>2833 | 6.9<br>[5.90, 7.90]<br>10939 | 6<br>[5.30, 6.90]<br>1585 | 6.1<br>[5.40, 7.20]<br>9525 |
| HBA1C | Glycated hemoglobin | mmol/mol | 38<br>[32.50, 40.50]<br>43 | 39<br>[35.00, 44.00]<br>197 | 42.5<br>[36.00, 55.00]<br>76 | 39<br>[35.02, 44.00]<br>1046 | 42.5<br>[37.00, 47.00]<br>32 | 40<br>[35.00, 48.00]<br>328 |
| HCO3 | Hydrogen carbonate | mmol/L | 24.9<br>[21.90, 27.40]<br>76 | 25.15<br>[22.82, 28.19]<br>148 | 23.4<br>[21.22, 25.92]<br>100 | 23.8<br>[22.30, 25.50]<br>526 | 24.3<br>[23.15, 25.40]<br>39 | 24.05<br>[21.58, 26.50]<br>256 |
| HDL | High density lipoprotein cholesterol | mmol/L | 1.02<br>[0.85, 1.40]<br>28 | 1.17<br>[0.88, 1.48]<br>140 | 1.01<br>[0.67, 1.35]<br>70 | 1.31<br>[1.04, 1.67]<br>972 | 1.05<br>[0.84, 1.47]<br>26 | 1.04<br>[0.81, 1.35]<br>235 |
| HR | Heart rate | $s^{-1}$ | 88<br>[77.01, 100.54]<br>1178 | 85.91<br>[74.00, 98.35]<br>4730 | 89.5<br>[78.50, 101.87]<br>2741 | 81.73<br>[71.50, 93.50]<br>10145 | 85.81<br>[75.35, 95.67]<br>1542 | 86.67<br>[75.14, 99.00]<br>9369 |
| INR | International normalized ratio blood test | ratio | 1.1<br>[1.00, 1.27]<br>795 | 1.1<br>[1.00, 1.30]<br>2632 | 1.2<br>[1.00, 1.30]<br>1272 | 1<br>[1.00, 1.20]<br>4269 | 1.1<br>[1.00, 1.20]<br>522 | 1.1<br>[1.00, 1.30]<br>5128 |
| JERN | Iron | µmol/L | 5.5<br>[4.00, 9.75]<br>30 | 7<br>[4.00, 14.75]<br>130 | 7<br>[4.00, 13.00]<br>61 | 8<br>[5.43, 14.00]<br>370 | 6<br>[4.00, 8.00]<br>41 | 6<br>[4.00, 10.00]<br>339 |
| K | Potassium | mmol/L | 4<br>[3.70, 4.30]<br>779 | 3.9<br>[3.60, 4.20]<br>3609 | 3.94<br>[3.60, 4.30]<br>2366 | 3.9<br>[3.60, 4.24]<br>9828 | 3.9<br>[3.50, 4.20]<br>1451 | 3.9<br>[3.60, 4.20]<br>7383 |
| KBGLU | Capillary blood glucose | mmol/L | 6.7<br>[5.75, 17.50]<br>15 | 7.7<br>[5.80, 10.02]<br>51 | 9<br>[6.30, 11.62]<br>32 | 6.3<br>[5.65, 8.24]<br>154 | 8.03<br>[7.03, 9.35]<br>20 | 8.3<br>[6.40, 10.61]<br>128 |
| LDH | Lactate dehydrogenase | U/L | 295<br>[205.00, 452.00]<br>625 | 259<br>[195.00, 367.00]<br>2027 | 240<br>[182.00, 359.00]<br>1233 | 221<br>[182.00, 286.00]<br>4010 | 237<br>[189.00, 314.00]<br>752 | 255<br>[192.00, 374.00]<br>3129 |
| LDL | Low density lipoprotein cholesterol | mmol/L | 2.55<br>[1.80, 3.55]<br>20 | 2<br>[1.30, 3.00]<br>85 | 2.1<br>[1.00, 2.50]<br>45 | 2.3<br>[1.70, 3.20]<br>477 | 2.4<br>[2.00, 3.68]<br>18 | 1.8<br>[1.10, 2.50]<br>153 |
| LEU | Leukocyte count | $10^9$/L | 10.9<br>[7.60, 14.83]<br>1176 | 10.7<br>[7.50, 14.60]<br>4597 | 8.5<br>[4.10, 13.00]<br>2690 | 9.1<br>[6.70, 12.30]<br>9946 | 9.4<br>[6.20, 13.00]<br>1521 | 10.5<br>[7.40, 14.60]<br>8932 |
| LYMFO | Lymphocyte count | $10^9$/L | 1.23<br>[0.71, 1.81]<br>509 | 1.19<br>[0.80, 1.73]<br>2399 | 0.95<br>[0.51, 1.50]<br>910 | 1.37<br>[0.93, 1.91]<br>5712 | 1.23<br>[0.76, 1.80]<br>596 | 1.22<br>[0.80, 1.81]<br>3236 |

| Variable | Description | Unit | Airways | PSAirways | BSI | UTI | PSUTI | Other |
|---|---|---|---|---|---|---|---|---|
| MBAS | Minimal biochemical abnormality score | % | 40 [40.00, 60.00] 1228 | 40 [20.00, 60.00] 4910 | 40 [20.00, 60.00] 2968 | 40 [20.00, 40.00] 11430 | 40 [40.00, 60.00] 1622 | 40 [20.00, 60.00] 9922 |
| MCH | Mean corpuscular hemoglobin | fmol | 1.9 [1.80, 2.00] 86 | 1.9 [1.80, 2.00] 430 | 1.9 [1.80, 2.00] 160 | 1.9 [1.80, 2.00] 1478 | 1.9 [1.80, 1.90] 126 | 1.8 [1.70, 1.90] 611 |
| MCHC | Mean corpuscular hemoglobin concentration | mmol/L | 20.25 [19.70, 21.00] 90 | 20.3 [19.60, 20.80] 440 | 20.4 [19.50, 21.10] 157 | 20.4 [19.80, 20.90] 1525 | 20.1 [19.40, 20.65] 127 | 20.1 [19.60, 20.70] 640 |
| MG | Magnesium | mmol/L | 0.88 [0.78, 0.96] 546 | 0.86 [0.77, 0.95] 1705 | 0.82 [0.72, 0.91] 952 | 0.84 [0.76, 0.92] 2436 | 0.82 [0.73, 0.94] 415 | 0.84 [0.74, 0.94] 2941 |
| MONO | Monocyte count | $10^9$/L | 0.7 [0.47, 0.96] 509 | 0.8 [0.57, 1.09] 2400 | 0.67 [0.38, 1.00] 911 | 0.7 [0.50, 0.97] 5713 | 0.73 [0.50, 1.02] 596 | 0.75 [0.50, 1.06] 3236 |
| NA | Sodium | mmol/L | 139 [136.00, 142.00] 791 | 139 [136.00, 142.00] 3651 | 137 [134.00, 140.00] 2384 | 138 [135.00, 141.00] 9945 | 138 [135.00, 141.00] 1458 | 138 [135.00, 140.00] 7493 |
| NEUTRO | Neutrophil count | $10^9$/L | 7.9 [5.47, 11.00] 495 | 7.85 [5.50, 11.00] 2346 | 7.13 [4.31, 11.00] 889 | 6.42 [4.48, 9.14] 5573 | 6.75 [4.56, 9.94] 573 | 7.58 [5.11, 11.00] 3081 |
| O2Therapy | Oxygen therapy | L/min | 35.15 [3.50, 50.87] 1113 | 25.43 [0.75, 40.73] 4348 | 0 [0.00, 25.86] 2104 | 0 [0.00, 2.67] 7918 | 0 [0.00, 2.00] 1245 | 1 [0.00, 34.21] 7865 |
| PHOS | Phosphate | mmol/L | 1.12 [0.88, 1.44] 567 | 1.06 [0.80, 1.41] 2043 | 1.09 [0.84, 1.42] 1103 | 1.08 [0.87, 1.32] 3307 | 1.14 [0.89, 1.59] 625 | 1.09 [0.83, 1.45] 3314 |
| PROCAL | Procalcitonin | μg/L | 0.66 [0.22, 2.59] 247 | 0.98 [0.30, 2.93] 594 | 0.81 [0.27, 2.94] 243 | 0.29 [0.10, 0.69] 344 | 1.17 [0.33, 2.48] 73 | 1.29 [0.34, 4.72] 1018 |
| RespFreq | Respiration frequency | $min^{-1}$ | 16.75 [13.32, 20.38] 1153 | 17 [14.00, 20.00] 4590 | 17.33 [16.00, 20.00] 2577 | 16.86 [15.50, 18.75] 9517 | 17.03 [16.00, 19.00] 1471 | 17 [15.31, 19.50] 8957 |
| Saturation | Blood oxygen saturation | % | 96.37 [94.72, 97.80] 1171 | 96.73 [95.13, 98.00] 4660 | 97.5 [96.17, 99.00] 2631 | 97.5 [96.00, 98.67] 9745 | 97.33 [96.00, 98.50] 1491 | 97.27 [96.00, 98.56] 9153 |
| SystolicBP | Systolic blood pressure | mmHg | 119.32 [106.00, 132.82] 1158 | 123.84 [110.80, 138.46] 4684 | 122 [107.33, 136.50] 2717 | 128.87 [115.25, 143.92] 10028 | 126 [114.00, 140.62] 1540 | 123.13 [109.42, 137.72] 9236 |
| TRANSJBG | Transferrin saturation | Interval [0, 1] | 0.13 [0.08, 0.18] 13 | 0.19 [0.12, 0.29] 81 | 0.15 [0.09, 0.26] 53 | 0.17 [0.11, 0.26] 309 | 0.13 [0.10, 0.19] 36 | 0.14 [0.10, 0.23] 271 |
| TRIG | Triglyceride | mmol/L | 2.17 [1.38, 2.79] 96 | 1.72 [1.17, 2.77] 312 | 1.73 [1.19, 3.11] 201 | 1.44 [1.06, 2.06] 1157 | 1.84 [1.19, 2.35] 59 | 1.71 [1.17, 2.84] 615 |
| TSH | Thyroid stimulating hormone | $10^{-3}$ IU/L | 2.26 [1.25, 4.77] 42 | 1.75 [0.94, 3.15] 210 | 1.63 [0.67, 2.87] 77 | 1.44 [0.81, 2.46] 1181 | 2.2 [1.20, 3.25] 43 | 1.45 [0.78, 2.35] 341 |
| Temperature | Temperature | °C | 37.15 [36.66, 37.64] 1145 | 37.15 [36.60, 37.67] 4607 | 37.1 [36.60, 37.75] 2650 | 36.95 [36.55, 37.41] 9624 | 36.9 [36.50, 37.40] 1498 | 37.03 [36.60, 37.51] 9082 |

| Variable | Description | Unit | Airways | PSAirways | BSI | UTI | PSUTI | Other |
|---|---|---|---|---|---|---|---|---|
| Weight | Weight | Kg | 80<br>[63.88, 95.00]<br>599 | 77.6<br>[62.18, 89.50]<br>2268 | 72.8<br>[57.50, 86.60]<br>1329 | 71.33<br>[58.57, 84.25]<br>4555 | 72.75<br>[62.48, 86.30]<br>724 | 76.3<br>[62.31, 88.80]<br>4554 |
| age | Age | Years | 59<br>[44.00, 69.00]<br>1228 | 65<br>[51.00, 74.00]<br>4910 | 62<br>[42.00, 71.00]<br>2968 | 67<br>[53.00, 76.00]<br>11430 | 66<br>[55.00, 74.00]<br>1622 | 64<br>[49.00, 73.00]<br>9920 |
| eGFR | Estimated glomerular filtration rate | ml/min | 77<br>[45.00, 90.00]<br>1061 | 79<br>[46.00, 90.00]<br>4379 | 71<br>[38.00, 90.00]<br>2452 | 80<br>[50.00, 90.00]<br>9870 | 63<br>[29.00, 90.00]<br>1507 | 80<br>[43.00, 90.00]<br>8363 |
| Sex | Sex | Binary | Female 449<br>Male 779 | Female 1577<br>Male 3333 | Female 1194<br>Male 1774 | Female 7361<br>Male 4069 | Female 901<br>Male 721 | Female 4131<br>Male 5791 |
